# Vitamin D supplementation to prevent acute respiratory infections: systematic review and meta-analysis of stratified aggregate data

**DOI:** 10.1101/2024.09.18.24313866

**Authors:** David A Jolliffe, Carlos A Camargo, John D Sluyter, Mary Aglipay, John F Aloia, Peter Bergman, Heike A. Bischoff-Ferrari, Arturo Borzutzky, Vadim Y Bubes, Camilla T Damsgaard, Francine Ducharme, Gal Dubnov-Raz, Susanna Esposito, Davaasambuu Ganmaa, Clare Gilham, Adit A Ginde, Inbal Golan-Tripto, Emma C Goodall, Cameron C Grant, Christopher J Griffiths, Anna Maria Hibbs, Wim Janssens, Anuradha Vaman Khadilkar, Ilkka Laaksi, Margaret T Lee, Mark Loeb, Jonathon L Maguire, Paweł Majak, Semira Manaseki-Holland, JoAnn E Manson, David T Mauger, David R Murdoch, Akio Nakashima, Rachel E Neale, Hai Pham, Christine Rake, Judy R Rees, Jenni Rosendahl, Robert Scragg, Dheeraj Shah, Yoshiki Shimizu, Steve Simpson-Yap, Geeta Trilok Kumar, Mitsuyoshi Urashima, Adrian R Martineau

## Abstract

**Background:** A 2021 meta-analysis of 37 randomised controlled trials (RCTs) of vitamin D supplementation for prevention of acute respiratory infections (ARI) revealed a statistically significant protective effect of the intervention (odds ratio [OR] 0.92, 95% confidence interval [CI] 0.86 to 0.99). Since then, 6 eligible RCTs have completed, including one large trial (n=15,804).

**Methods:** Updated systematic review and meta-analysis of data from RCTs of vitamin D for ARI prevention using a random effects model. Sub-group analyses were done to determine whether effects of vitamin D on risk of ARI varied according to baseline 25-hydroxyvitamin D (25[OH]D) concentration, dosing regimen or age. We searched MEDLINE, EMBASE, the Cochrane Central Register of Controlled Trials, Web of Science and the ClinicalTrials.gov were searched between May 2020 (previous search) and April 2024. No language restrictions were imposed. Double-blind RCTs supplementing vitamin D for any duration, with placebo or low-dose vitamin D control, were eligible if approved by Research Ethics Committee and if ARI incidence was collected prospectively and pre-specified as an efficacy outcome. Aggregate data, stratified by baseline 25(OH)D concentration and age, were obtained from study authors. The study was registered with PROSPERO (no. CRD42024527191).

**Findings:** We identified 6 new RCTs (19,337 participants). Data were obtained for 16,086 (83.2%) participants in 3 new RCTs and combined with data from 48,488 participants in 43 previously identified RCTs. For the primary comparison of any vitamin D vs. placebo, the intervention did not significantly affect overall ARI risk (OR 0.94, 95% CI 0.88 to 1.00, P=0.057; 40 studies; I^2^ 26.4%). Pre-specified subgroup analysis did not reveal evidence of effect modification by age, baseline vitamin D status, or dosing regimen. Vitamin D did not influence the proportion of participants experiencing at least one serious adverse event (OR 0.96, 95% CI 0.90 to 1.04; 38 studies; I^2^ 0.0%). A funnel plot showed left-sided asymmetry (P=0.002, Egger’s test).

**Interpretation:** This updated meta-analysis yielded a similar point estimate for the overall effect of vitamin D supplementation on ARI risk to that obtained previously, but the 95% CI for this effect estimate now spans 1.00, indicating no statistically significant protection.

**Funding:** None

**Research in context:** *Evidence before this study:* We searched MEDLINE, EMBASE, the Cochrane Central Register of Controlled Trials (CENTRAL), Web of Science and the ClinicalTrials.gov registry from 1^st^ May 2020 (date of our previous search) to 30^th^ April 2024 for randomised Controlled Trials and meta-analyses of randomised Controlled Trials evaluating effectiveness of vitamin D supplementation for the prevention of acute respiratory infections. Our previous meta-analysis of 43 randomised Controlled Trials (RCTs) of vitamin D supplementation for prevention of acute respiratory infections (ARI) conducted in 2021 revealed a statistically significant protective effect of the intervention (OR 0.92, 95% CI 0.86 to 0.99). A further 6 eligible RCTs, contributing data from 19,337 participants have now completed, including one large trial (n=15,804).

*Added value of this study:* Our meta-analysis of aggregate data from 64,086 participants in 46 randomised controlled trials, stratified by baseline 25(OH)D concentration and age, provides an updated estimate of the protective effects of vitamin D against acute respiratory infection overall (OR 0.94, 95% CI 0.88 to 1.00), and in sub-groups defined by baseline vitamin D status, age, and dosing frequency, amount and duration.

*Implications of all the available evidence:* Updated meta-analysis including the latest available RCT data shows no statistically significant protective effect of vitamin D supplementation against ARI, either overall or in sub-group analyses.

## Introduction

The contribution of acute respiratory infections (ARI) to global morbidity and mortality, with consequent strain on healthcare systems, remains an ongoing problem in the post-COVID pandemic era. Evidence indicating that vitamin D supplementation could reduce risk of ARI arises from laboratory studies which show that vitamin D metabolites support innate immune responses to respiratory viruses,^1^ together with observational studies reporting independent associations of low circulating levels of 25-hydroxyvitamin D (25[OH]D, the widely accepted biomarker of vitamin D status) and increased risk of ARI.^2,3^

Randomised controlled trials (RCTs) of vitamin D for the prevention of ARI have produced heterogeneous results, with some showing protection, and others reporting null findings. We previously meta-analysed aggregate data from 48,488 participants in 43 RCTs,^4–46^ and showed a modest protective overall effect that was stronger in trials which administered vitamin D in daily, that used doses of 400-1000 IU/day, were up to 12 months in length, and that were conducted among participants aged 1.00 to 15.99 years at enrolment.^47^ Since the date of our previous literature search (May 2020), 6 RCTs with 19,337 participants fulfilling the same eligibility criteria have been completed. We therefore sought data from these recent studies for inclusion in an updated meta-analysis of stratified aggregate data (trial-level, stratified by baseline vitamin D status and age) to determine whether vitamin D reduced ARI risk overall, and to evaluate whether effects of vitamin D on ARI risk varied according to baseline 25(OH)D concentration, dosing regimen (frequency, dose size, and trial duration) or age at enrolment.

## Methods

### Protocol, Registration and Ethical Approvals

Methods were pre-specified in a protocol that was registered with the PROSPERO International Prospective Register of Systematic Reviews (<u>click here</u>). Details of Research Ethics Committee approvals to conduct this study are included in Supplementary Material (Section 4).

### Eligibility Criteria

Double-blind randomised controlled trials of supplementation with vitamin D_3_, vitamin D_2_ or 25(OH)D of any duration, with a placebo or blinded low-dose vitamin D control, were eligible for inclusion if they had been approved by a Research Ethics Committee and if data on incidence of ARI were collected prospectively and pre-specified as an efficacy outcome. The latter requirement was imposed to minimise misclassification bias (prospectively designed instruments to capture ARI events were deemed more likely to be sensitive and specific for this outcome). Studies reporting results of long-term follow-up of primary RCTs were excluded.

### Study Identification and Selection

Two investigators (ARM and DAJ) searched MEDLINE, EMBASE, the Cochrane Central Register of Controlled Trials (CENTRAL), Web of Science and the ClinicalTrials.gov registry using the electronic search strategies described in the Supplementary Material (Section 1). Searches were regularly updated up to and including 1^st^ April 2024. No language restrictions were imposed. These searches were supplemented by searching review articles and reference lists of trial publications. Collaborators were asked if they knew of any additional eligible RCTs.

### Data Collection Process and Definition of outcomes

Details of the data collection process can be found in Supplementary Material (Section 2). The primary outcome of the meta-analysis was the proportion of participants experiencing one or more ARI, with the definition of ARI encompassing events classified as URI, LRI and ARI of unclassified location (i.e. infection of the upper and/or lower respiratory tract). Secondary outcomes were: incidence of URI and LRI, analysed separately; incidence of Emergency Department attendance and/or hospital admission for ARI; death due to ARI or respiratory failure; use of antibiotics to treat an ARI; absence from work or school due to ARI; incidence of serious adverse events; death due to any cause; and incidence of potential adverse reactions to vitamin D (hypercalcaemia and renal stones).

### Risk of Bias Assessment for Individual Studies

We used the Cochrane Collaboration Risk of Bias tool^48^ to assess the following variables: sequence generation, allocation concealment, blinding of participants, personnel and outcome assessors, completeness of outcome data, evidence of selective outcome reporting, and other potential threats to validity. Study quality was assessed independently by two investigators (ARM and DAJ), except for the six trials for which DAJ and/or ARM were investigators, which were assessed by CAC and JDS. Discrepancies were resolved by consensus.

### Synthesis Methods

Data were analysed by DAJ; results were checked and verified by JDS. Our meta-analysis approach followed published guidelines.^49^ The primary comparison was of participants randomised to any vitamin D supplement vs. placebo: this was performed for all of the outcomes listed above. For trials that included higher-dose, lower-dose and placebo arms, data from higher-dose and lower-dose arms were pooled for analysis of the primary comparison. A secondary comparison of participants randomised to higher vs. lower doses of vitamin D was performed for the primary outcome only.

The log odds ratio and its standard error were calculated for each outcome within each trial from the proportion of participants experiencing one or more events in the intervention vs. control arm. For trials where randomisation was stratified by study site, proportions were corrected for clustering using published methods.^50^ Proportions were then meta-analysed in a random effects model using the Metan package^51^ within STATA IC v14.2 to obtain a pooled odds ratio with a 95% confidence interval (CI) and a measure of heterogeneity summarized by the I^2^ statistic and its corresponding P value.

### Exploration of variation in effects

To explore reasons for heterogeneity of effect of the intervention between trials we performed a stratified analysis according to baseline vitamin D status (serum 25[OH]D <25 *vs.* 25-49.9 *vs.* 50-74.9 *vs.* ≥75 nmol/L) and according to age at baseline (<1.00 *vs.* 1.00-15.99 *vs.* 16.00-64.99 *vs.* ≥65.00 years). We also conducted sub-group analyses according to vitamin D dosing regimen (administration of daily *vs.* weekly *vs.* monthly or less frequent doses), dose size (daily equivalent <400 IU *vs.* 400-1000 IU *vs.* 1001-2000 IU *vs.* >2,000 IU), trial duration (≤12 months *vs.* >12 months) and presence of airway disease (trial restricted to participants with asthma *vs.* those restricted to participants with COPD *vs.* those in which participants without airway disease were eligible). The thresholds for baseline 25(OH)D concentration used in sub-group analyses were selected *a priori* on the basis that they represent cut-offs that are commonly used to distinguish profound vitamin D deficiency (<25 nmol/L), moderate vitamin D deficiency (25-49.9 nmol/L) and potentially sub-optimal vitamin D status (50-74.9 nmol/L).^52^

To investigate factors associated with heterogeneity of effect between statistically significant subgroups of trials, we performed multivariable meta-regression analysis on trial-level characteristics, the full details of which is described in Supplementary Material (Section 5).

### Quality Assessment Across Studies

For the primary analysis, the likelihood of publication bias was investigated through the construction of a contour-enhanced funnel plot.^53^ We used the five GRADE considerations (study limitations, consistency of effect, imprecision, indirectness and publication bias)^54^ to assess the quality of the body of evidence contributing to analyses of the primary efficacy outcome and major secondary outcomes of our meta-analysis.

### Sensitivity analyses

We conducted two exploratory sensitivity analyses for the primary comparison of the primary outcome: one excluded RCTs where risk of bias was assessed as being unclear, and the other excluded RCTs in which incidence of ARI was not the primary or co-primary outcome.

### Exploratory analyses

Due to the relatively low level of heterogeneity between trials entering into the primary outcome model, we also estimated the overall primary outcome using a fixed effects model.

## Results

### Study selection and data obtained

The study selection process is illustrated in Figure 1. Our updated search (studies registered from 2^nd^ May 2020 to 1^st^ April 2024) identified a total of 1,107 studies that were assessed for eligibility, of which six studies with a total of 19,337 randomised participants fulfilled eligibility criteria. Studies for which full text was reviewed prior to exclusion due to ineligibility are listed in Table S1. All six of the eligible studies identified compared effects of a single vitamin D regimen vs. placebo only. Data for the primary outcome (proportion of participants with one or more ARI) were obtained for 15,598 (98.1%) of 16,085 participants in 3 studies^14,55,56^ and were added to our database of 43 previously identified eligible studies, described elsewhere,^47^ bringing the total number of participants contributing data to analysis of our primary outcome to 64,086 out of 65,504 (97.8%) participants in 46 studies.

**Figure 1:**
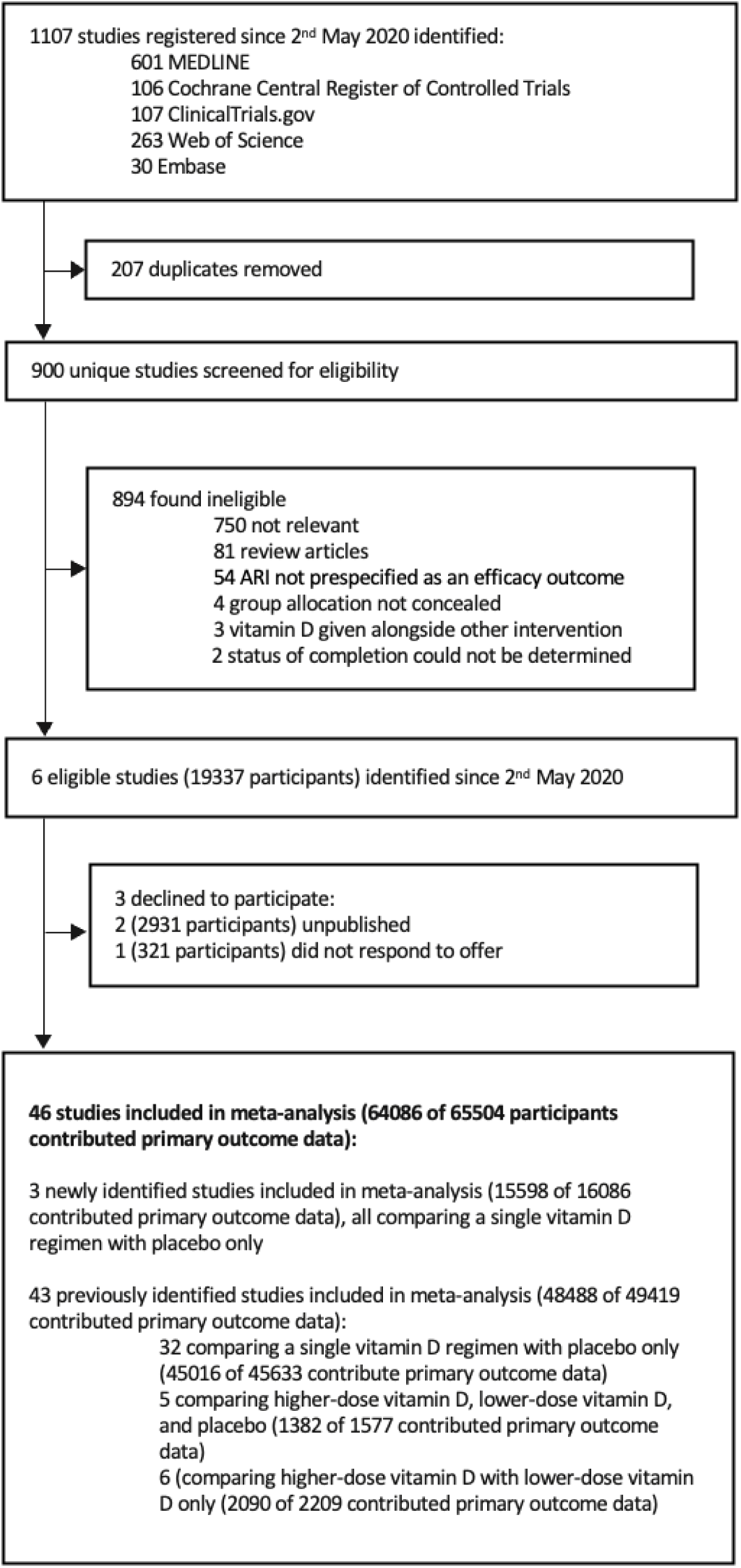
Flow chart of study selection

### Study and participant characteristics

Characteristics of the 46 studies contributing data to this meta-analysis are presented in Table 1. Trials were conducted in 24 different countries on 5 continents, and enrolled participants of both sexes from birth to 100 years of age. Baseline serum 25(OH)D concentrations were determined in 38 of 46 trials: mean baseline 25(OH)D concentration ranged from 18.9 to 90.9 nmol/L (to convert to ng/mL, divide by 2.496). Forty-five studies administered oral vitamin D_3_ to participants in the intervention arm, while 1 study administered oral 25(OH)D. Vitamin D was given as monthly to 3-monthly bolus doses in 13 studies; as weekly doses in 7 studies; as daily doses in 24 studies; and as a combination of bolus and daily doses in 2 studies. Trial duration ranged from 7 weeks to 5 years. Incidence of ARI was primary or co-primary outcome for 25 studies, and a secondary outcome for 21 studies.

**Table 1:**
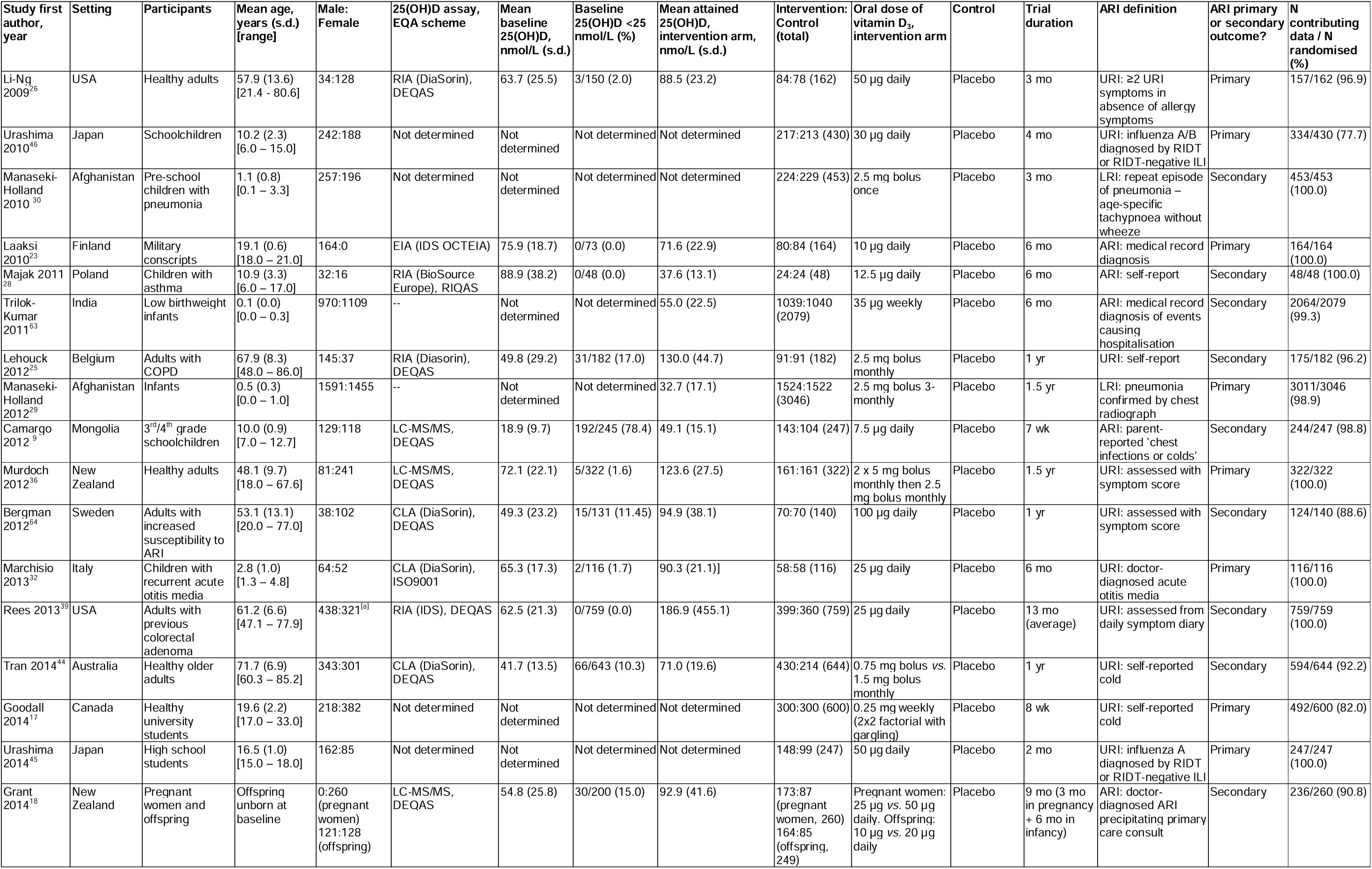
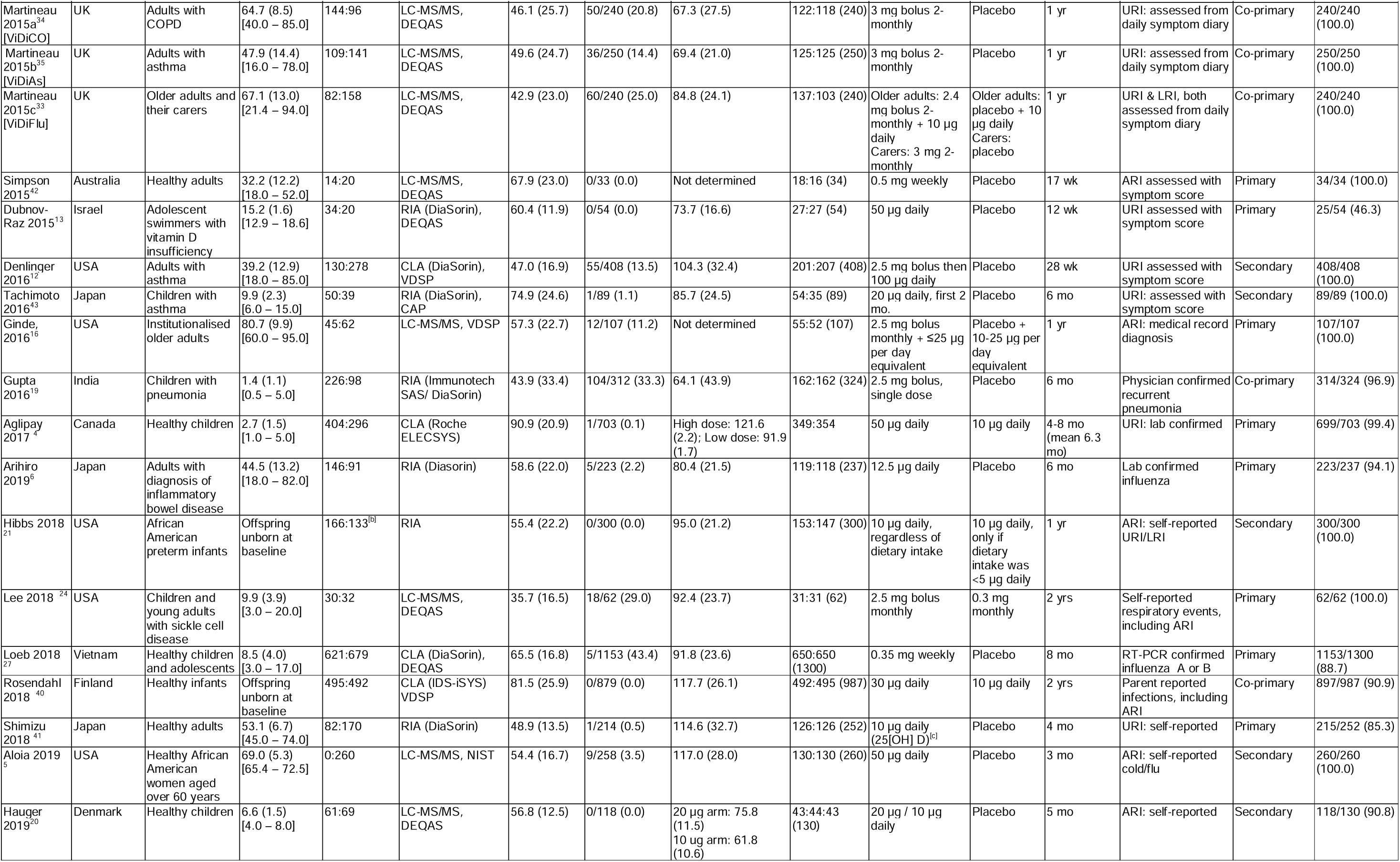
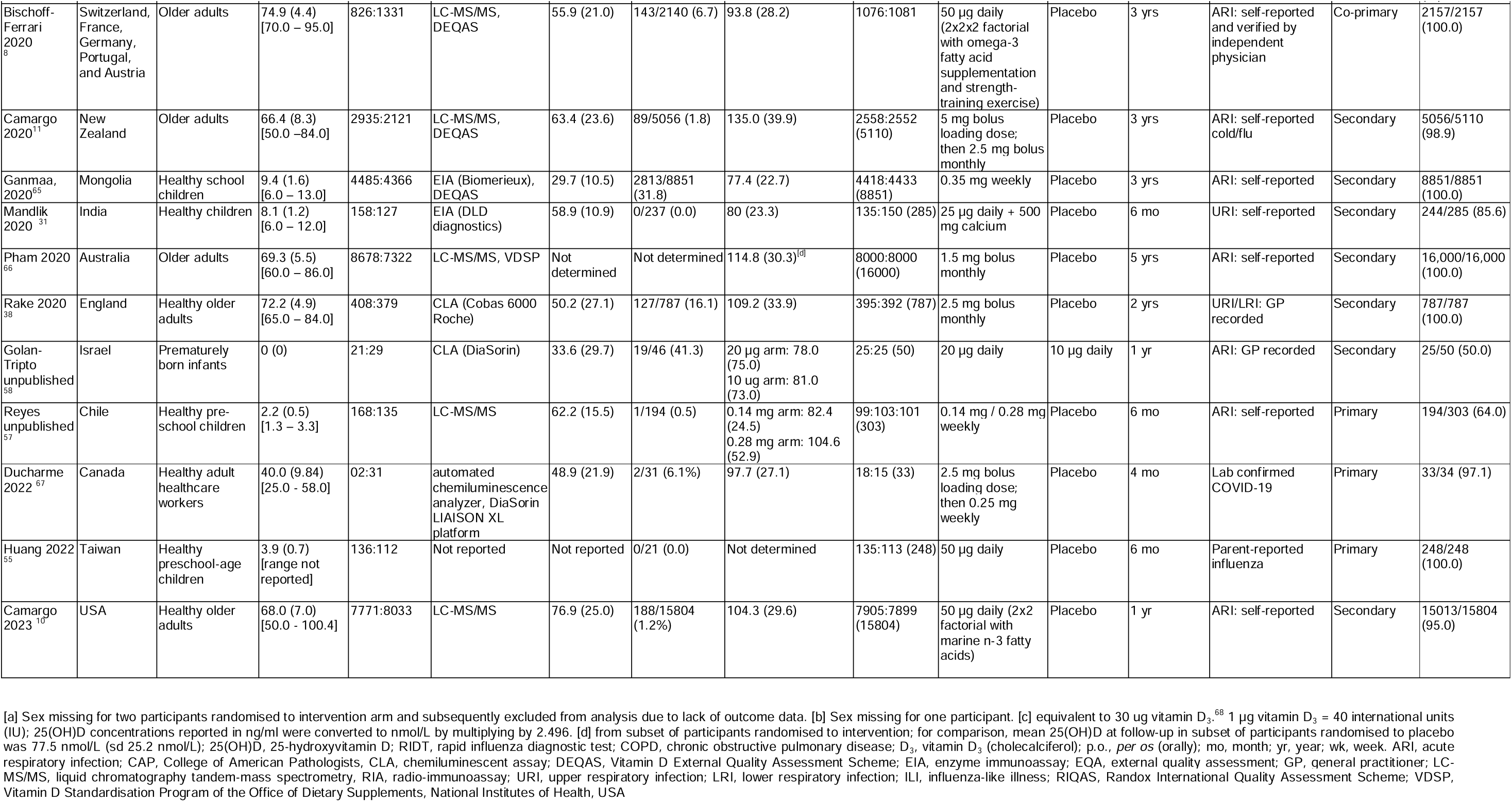
Characteristics of trials and their participants.

### Risk of Bias Within Studies

Details of the risk of bias assessment are provided in supplementary Table S2. Five trials were assessed as being at unclear risk of bias due to high loss to follow-up. In the trial by Laaksi and colleagues,^23^ 37% of randomised participants were lost to follow-up. In the trial by Dubnov-Raz and colleagues,^13^ 52% of participants did not complete all symptom questionnaires. In the unpublished trial by Reyes and colleagues,^57^ loss to follow-up ranged from 33% to 37% across the three study arms, and in the unpublished trial by Golan-Tripto and colleagues,^58^ 50% of participants were lost to follow-up. Finally, in the trial by Huang and colleagues,^55^ we detected uncertainty around blinding of outcome assessment within the study team; methodology for dealing with incomplete data, and selective outcome reporting, which we were unable to resolve with the authors. All other trials were assessed as being at low risk of bias for all seven aspects assessed.

### Overall Results, Primary Outcome

For the primary comparison of any vitamin D supplement vs. placebo control, supplementation did not result in a statistically significant reduction in the proportion of participants experiencing at least one ARI (odds ratio [OR] 0.94, 95% CI 0.88 to 1.00, P=0.57; 61,589 participants in 40 studies; Figure 2, Table 2; Cates Plot, Figure S1). Between-trial heterogeneity was modest: I^2^ = 26.4% (P for heterogeneity 0.07).

**Figure 2:**
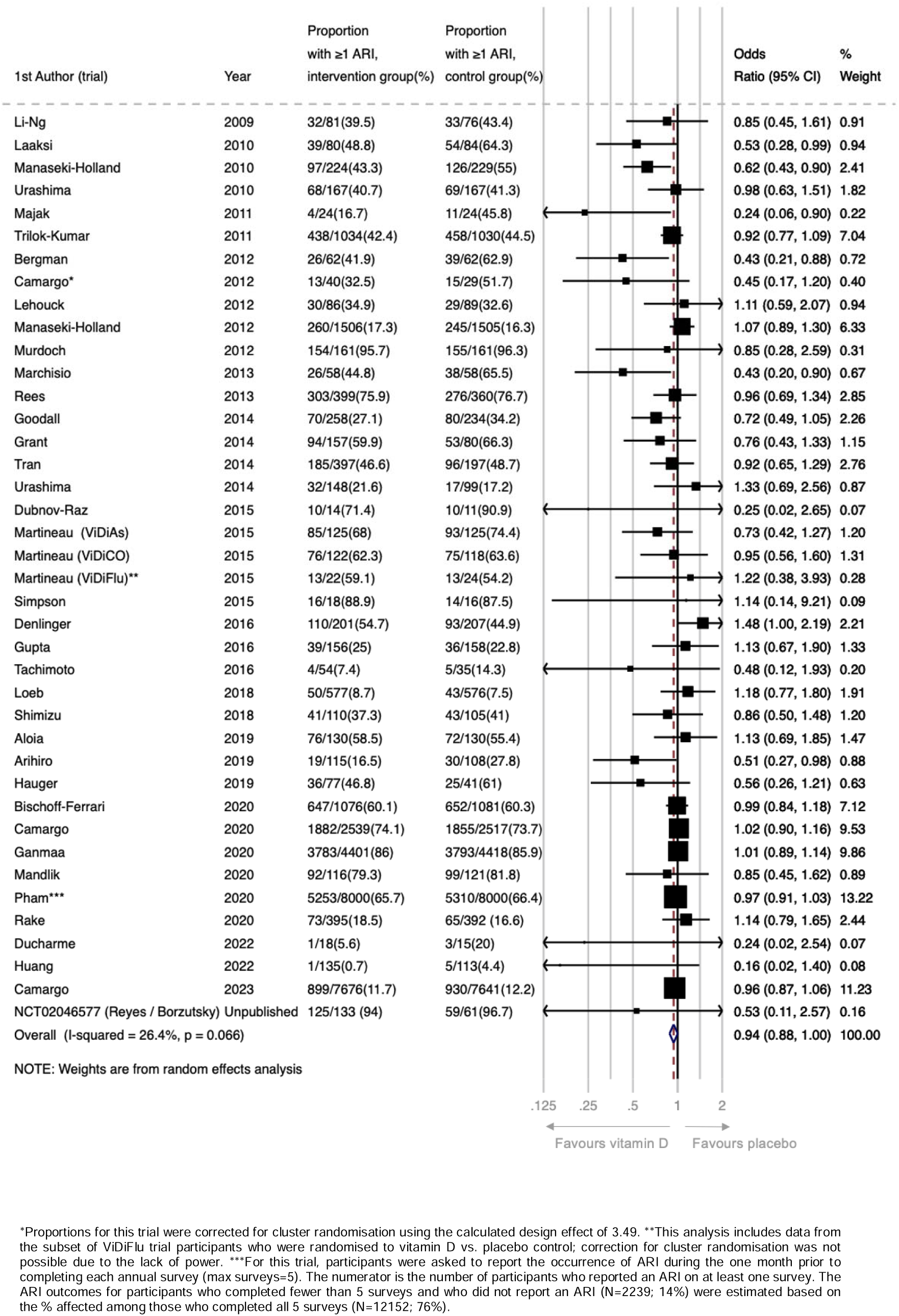
Forest plot of placebo-controlled RCTs reporting proportion of participants experiencing 1 or more acute respiratory infection.

**Table 2:** Placebo controlled RCTs: Proportion of participants experiencing at least one acute respiratory infection, overall and stratified by potential effect-modifiers

| Potential effect-modifier | No of trials | Proportion with ≥1 ARI, intervention group (%) | Proportion with ≥1 ARI, control group (%) | Odds ratio (95% CI) | I<sup>2</sup> % | P for heterogeneity |
| --- | --- | --- | --- | --- | --- | --- |
| **Overall** | 40 | 15202/31092 (48.9) | 15117/30497 (49.6) | 0.94 (0.88 to 1.002) | 26.4 | 0.07 |
| **Baseline 25(OH)D, nmol/L<sup>[a]</sup>** |  |  |  |  |  |  |
| <25 | 22 | 1387/1893 (73.3) | 1408/1913 (73.6) | 0.98 (0.80 to 1.20) | 3.6 | 0.41 |
| 25 – 49.9 | 31 | 3783/5849 (64.7) | 3707/5769 (64.3) | 1.03 (0.94 to 1.13) | 0.0 | 0.55 |
| 50 – 74.9 | 32 | 2237/5749 (38.9) | 2142/5465 (39.2) | 0.90 (0.80 to 1.02) | 8.7 | 0.33 |
| ≥75 | 28 | 1530/6045 (25.3) | 1503/5899 (25.5) | 0.97 (0.87 to 1.07) | 0.0 | 0.83 |
| **Dosing frequency** |  |  |  |  |  |  |
| Daily | 21 | 2572/10920 (23.6) | 2569/10632 (24.2) | 0.84 (0.73 to 0.97) | 44.8 | 0.01 |
| Weekly | 7 | 4483/6439 (69.6) | 4450/6350 (70.1) | 0.97 (0.88 to 1.06) | 0.0 | 0.44 |
| Monthly or less frequently | 12 | 8147/13733 (59.3) | 8098/13515 (59.9) | 0.98 (0.93 to 1.03) | 0.0 | 0.57 |
| **Daily dose equivalent, IU<sup>[b]</sup>** |  |  |  |  |  |  |
| <400 | 2 | 451/1074 (42.0) | 473/1059 (44.7) | 0.76 (0.41 to 1.41) | 49.0 | 0.16 |
| 400-1000 | 10 | 656/1236 (53.1) | 627/1069 (58.7) | 0.70 (0.55 to 0.89) | 31.2 | 0.16 |
| 1001-2000 | 19 | 11494/24790 (46.4) | 11612/24667 (47.1) | 0.97 (0.92 to 1.01) | 1.6 | 0.44 |
| >2000 | 7 | 2291/3462 (66.2) | 2250/3444 (65.3) | 1.05 (0.84 to 1.31) | 37.1 | 0.15 |
| **Trial duration, months** |  |  |  |  |  |  |
| ≤12 | 32 | 2847/12615 (22.6) | 2766/12063 (23.0) | 0.85 (0.76 to 0.95) | 32.7 | 0.04 |
| >12 | 8 | 12355/18477 (66.9) | 12351/18434 (67.0) | 0.99 (0.95 to 1.04) | 0.0 | 0.95 |
| **Age, yrs<sup>[a]</sup>** |  |  |  |  |  |  |
| <1.00 | 5 | 875/2901 (30.2) | 839/2796 (30.0) | 0.95 (0.82 to 1.10) | 18.7 | 0.30 |
| 1.00-15.99 | 16 | 4267/6028 (70.8) | 4271/5916 (72.2) | 0.74 (0.60 to 0.92) | 33.2 | 0.10 |
| 16.00-64.99<sup>[c]</sup> | 22 | 3138/4894 (64.1) | 3090/4742 (65.2) | 0.96 (0.85 to 1.08) | 12.5 | 0.29 |
| ≥65.00<sup>[c]</sup> | 17 | 6023/9665 (62.3) | 6004/9475 (63.4) | 0.96 (0.90 to 1.02) | 0.0 | 0.73 |
| **Airway disease** |  |  |  |  |  |  |
| Asthma only | 4 | 203/404 (50.2) | 202/391 (51.7) | 0.73 (0.36 to 1.49) | 71.7 | 0.01 |
| COPD only | 2 | 106/208 (51.0) | 104/207 (50.2) | 1.01 (0.68 to 1.51) | 0.0 | 0.71 |
| Unrestricted | 34 | 14893/30480 (48.9) | 14811/29899 (49.5) | 0.94 (0.89 to 1.00) | 26.4 | 0.14 |

ory
[b] Data from two trials that included higher-dose, lower-dose and placebo arms<sup>44,57</sup> are excluded from this sub-group analysis, since the higher-dose and lower-dose arms spanned the 1,000 IU/day cut-off, rendering them unclassifiable
[c] Age-stratified data for one trial<sup>10</sup> were unavailable
[a] The number of trials in each category for this variable adds up to more than 40, since this is a participant-level variable, i.e. some trials contributed data from participants who fell into more than one category

For the secondary comparison of higher-vs. lower-dose vitamin D, we observed no statistically significant difference in the proportion of participants with at least one ARI (OR 0.87, 95% CI 0.73 to 1.04; 3,047 participants in 11 studies; I^2^ 0.0%, P for heterogeneity 0.50; Figure S2).

### Sub-group Analyses, Primary Outcome

To investigate reasons for the observed heterogeneity of effect for the primary comparison of any vitamin D supplement vs. placebo control, we stratified this analysis by two participant-level factors (baseline vitamin D status and age) and by four trial-level factors (dose frequency, dose size, trial duration, and airway disease comorbidity). Results are presented in Table 2 and Figures S3-S8. No statistically significant effect of vitamin D was seen for participants with baseline 25(OH)D <25 nmol/L (OR 0.98, 95% CI 0.80 to 1.20; 3,806 participants in 22 studies), 25-49.9 nmol/L (OR 1.03, 95% CI 0.94 to 1.13; 11,618 participants in 31 studies), 50-74.9nmol/L (OR 0.90, 95% CI 0.80 to 1.02; 11,214 participants in 32 studies), or ≥75 nmol/L (OR 0.97, 95% CI 0.87 to 1.07; 11,815 participants in 28 studies; Figure S3). A statistically significant protective effect of vitamin D was seen for participants aged 1.00-15.99 years (OR 0.74, 95% CI 0.60 to 0.92; 11,944 participants in 16 studies), but not in participants aged <1 year (OR 0.95, 95% CI 0.82 to 1.10; 5,697 participants in 5 studies), 16.00-64.99 years (OR 0.96, 95% CI 0.85 to 1.08; 9,636 participants in 22 studies), or ≥65.00 years (OR 0.96, 95% CI 0.90 to 1.02; 19,140 participants in 17 studies; Figure S7). With regard to dosing frequency, a statistically significant protective effect was seen for trials where vitamin D was given daily (OR 0.84, 95% CI 0.73 to 0.97; 21,552 participants in 21 studies), but not for trials in which it was given weekly (OR 0.97, 95% CI 0.88 to 1.06; 12,789 participants in 7 studies), or monthly to 3-monthly (OR 0.98, 95% CI 0.93 to 1.03; 27,248 participants in 12 studies; Figure S4). Statistically significant protective effects of the intervention were also seen in trials where vitamin D was administered at daily equivalent doses of 400-1000 IU (OR 0.70, 95% CI 0.55 to 0.89; 2,305 participants in 10 studies), but not where the daily dose equivalent was <400 IU (OR 0.76, 95% CI 0.41 to 1.41; 2,133 participants in 2 studies), 1001-2000 IU (OR 0.97, 95% CI 0.92 to 1.01; 49,457 participants in 19 studies), or >2000 IU (OR 1.05, 95% CI 0.84 to 1.31; 6,906 participants in 7 studies; Figure S5). Statistically significant protective effects were also seen for trials with a duration of ≤12 months (OR 0.85, 95% CI 0.76 to 0.95; 24,678 participants in 32 studies) but not in those lasting >12 months (OR 0.99, 95% CI 0.95 to 1.04; 36,911 participants in 8 studies; Figure S6).

Statistically significant protective effects of vitamin D were not seen in trials that exclusively enrolled participants with asthma, or trials that exclusively enrolled participants with COPD, or trials that were not restricted to participants with either asthma or COPD (Figure S8).

### Multivariable Meta-Regression Analysis

Multivariable meta-regression analysis of trial-level sub-groups did not identify any statistically significant interactions (P values for interaction <0.05) between allocation to vitamin D vs. placebo and dose frequency, dose size, trial duration or participant age (Table S3).

### Secondary outcomes

Meta-analysis of secondary outcomes was performed for results of placebo-controlled trials only (i.e. not for RCTs that compared higher- vs. lower-dose vitamin D). Results are presented in Table 3. Overall, without consideration of participant- or trial-level factors, vitamin D supplementation did not have a statistically significant effect on the proportion of participants with one or more URI, LRI, hospitalisations or emergency department attendances for ARI, death due to ARI or respiratory failure, courses of antimicrobials for ARI, work/school absences due to ARI, serious adverse events of any cause, death due to any cause, or episodes of hypercalcaemia or renal stones.

**Table 3:** Placebo-controlled studies: Secondary outcomes

| Variables | No of trials | Proportion with $\geq 1$ event, intervention group (%) | Proportion with $\geq 1$ event, control group (%) | Odds ratio (95% CI) | I<sup>2</sup> % | P for heterogeneity |
| --- | --- | --- | --- | --- | --- | --- |
| **Efficacy outcomes** |  |  |  |  |  |  |
| Upper respiratory infection | 32 | 9396/22244 (42.2) | 9341/21734 (43.0) | 0.95 (0.91 to 1.01) | 4.6 | 0.39 |
| Lower respiratory infection | 16 | 4040/20915 (19.3) | 4049/20739 (19.5) | 0.99 (0.93 to 1.04) | 0.0 | 0.58 |
| Emergency department attendance and/or hospital admission due to ARI | 20 | 139/10981 (1.3) | 149/10865 (1.4) | 0.90 (0.71 to 1.14) | 0.0 | 1.00 |
| Death due to ARI or respiratory failure | 35 | 14/14706 (0.1) | 11/14154 (0.1) | 1.03 (0.61 to 1.75) | 0.0 | 1.00 |
| Use of antibiotics to treat an ARI | 15 | 2056/8656 (23.8) | 2109/8519 (24.8) | 0.93 (0.86 to 1.01) | 2.0 | 0.43 |
| Absence from work or school due to ARI | 11 | 378/1545 (24.5) | 364/1059 (34.4) | 0.91 (0.70 to 1.18) | 28.1 | 0.18 |
| **Safety outcomes** |  |  |  |  |  |  |
| Serious adverse event of any cause | 38 | 1579/22860 (6.9) | 1621/22321 (7.3) | 0.96 (0.90 to 1.04) | 0.0 | 1.00 |
| Death due to any cause | 37 | 438/22853 (1.9) | 397/22288 (1.8) | 1.09 (0.95 to 1.25) | 0.0 | 1.00 |
| Hypercalcaemia | 23 | 143/18275 (0.8) | 124/17899 (0.7) | 1.13 (0.89 to 1.44) | 0.0 | 1.00 |
| Renal stones | 23 | 415/20539 (2.0) | 401/20133 (2.0) | 1.03 (0.90 to 1.18) | 0.0 | 1.00 |

\* This analysis includes a subset of participants in the trial by Pham et al, who completed symptom diaries.

### Risk of bias across studies

A funnel plot for the proportion of participants experiencing at least one ARI (Figure S9) showed left-sided asymmetry, confirmed with an Egger’s regression test^59^ (P=0.002). This might reflect heterogeneity of effect across trials, or publication bias arising from omission of small trials showing non-protective effects of vitamin D from the meta-analysis.^60^ Given the latter possibility, the quality of the body of evidence contributing to analyses of the primary efficacy outcome and major secondary outcomes was downgraded to moderate (Table S4).

### Sensitivity Analyses

Results of exploratory sensitivity analyses are presented in Table S5. Meta-analysis of the proportion of participants in placebo-controlled trials experiencing at least one ARI, excluding 4 studies assessed as being at unclear risk of bias,^13,23,55,57^ did not reveal a statistically significant protective effect of any vitamin D supplementation (OR 0.95, 95% CI 0.90 to 1.01; 60,958 participants in 36 studies), consistent with the main analysis. Similarly, a sensitivity analysis for the same outcome, excluding 19 placebo-controlled trials that investigated ARI as a secondary outcome, did not show a statistically significant protective effect (OR 0.90, 95% CI 0.79 to 1.02; 9,975 participants in 21 studies).

### Exploratory analysis

Due to the relatively low level of between-trial heterogeneity (I^2^ = 26.4%) we analysed the primary outcome using a fixed effects model, which yielded a very similar effect estimate (P=0.047).

## Discussion

This update to our 2021 meta-analysis of RCTs of vitamin D supplementation for the prevention of ARI includes new primary outcome data from an additional 15,598 participants in 3 studies completed since May 2020, bringing the total number contributing data to 64,086 participants in 46 trials. The point estimate of the overall effect of vitamin D supplementation on ARI risk obtained in the current analysis (0.94) is similar to that yielded by our previous meta-analysis (0.92). However, in contrast to our previous work, ^47^ the 95% CI for this effect now spans 1.00. Although statistically significant protection was seen within some trial subsets (daily dosing trials, trials that administered 400-1000 IU/day, trials conducted for 12 months or less and trials in participants aged 1.00-15.99 years), meta-regression analysis did not yield formal evidence to suggest that effects of vitamin D were modified by any of these factors.

Heterogeneity of results from the current meta-analysis is somewhat lower than that obtained from our previous meta-analysis (I^2^ 26.4% vs. 35.6%, respectively). This difference suggests that greater confidence may be placed in the findings of the current analysis vs. our previous one. It is possible that the previous overall finding of a protective effect of any vitamin D supplement was driven by small study effects, as evidenced by left-sided asymmetry shown in the funnel plot (Figure S9).^60^

The current study has several strengths: it contains the latest aggregate RCT data available worldwide, including stratified data for subgroups of baseline vitamin D status and age, new data from a very large trial (n=15,804)^10^ and as yet unpublished data, which together represent 10.7% of the overall model weight. The larger sample size provides improved statistical power to perform subgroup analyses and interrogate heterogeneity of effects across trials. Nevertheless, formal demonstration of effect modification is challenging and will likely require even larger sample sizes.

Our work also has limitations: some trials did not respond to our invitation to contribute data for meta-analysis (Figure 1 and Table S1), at least one of which reported protective effects of vitamin D against ARI,^61^ therefore potentially biasing our results towards the null. We meta-analysed aggregate (trial-level) data, rather than individual participant data. However, we did contact authors to get unpublished estimates of effect that were stratified by pre-defined baseline 25(OH)D levels and age, harmonised across studies: thus, we were able to obtain accurate data for the major participant-level potential effect-modifiers of interest. As with our previous update to synthesis of this research question, there are still relatively few RCTs that have compared effects of lower- vs. higher-dose vitamin D. Paucity of data in this area limited our power for this secondary comparison. We lacked the data to investigate race/ethnicity and obesity as potential effect-modifiers. We also could not account for other factors that might influence the efficacy of vitamin D supplements for ARI prevention (e.g., taking the supplement with or without food, calcium intake, vitamin A status) or secular trends that might influence trial findings, such as the increased societal use of vitamin D supplements:^62^ concurrent use of supplements containing vitamin D by participants randomised to control would effectively render these as high- vs. low-dose trials and potentially drive results toward the null. Another potential limitation is illustrated by the funnel plot, which suggests that the overall effect size may have been over-estimated due to publication bias; we have attempted to mitigate this problem by inclusion of data from unpublished studies identified by searching clinicaltrials.gov where this was obtainable.

In summary, this updated meta-analysis of data from RCTs of any vitamin D supplementation for the prevention of ARI yielded a similar point estimate for the overall effect of vitamin D supplementation on ARI risk to that obtained previously, but the 95% CI for this effect now spans 1.00, indicating no statistically significant protection.

## Data Availability

the study dataset is available from.

## Acknowledgements

This study was conducted without external funding. ARM is supported by the United Kingdom Office for Students. The views expressed are those of the authors and not necessarily those of Barts Charity or the Office for Students. Sources of support for individual trials are detailed in Supplementary Material. We thank all the people who participated in primary randomised controlled trials, and the teams who conducted them.

## Author Contributions

DAJ and ARM wrote the study protocol and designed statistical analyses. DAJ, CAC and ARM assessed eligibility of studies for inclusion. DAJ, ARM, CAC and JDS performed risk of bias assessments. DAJ and ARM had access to, and verified, the underlying data from all original research articles. Statistical analyses were done by DAJ; results were checked and verified by JDS. DAJ and ARM wrote the first draft of the report. All authors revised it critically for important intellectual content, gave final approval of the version to be published, and agreed to be accountable for all aspects of the work in ensuring that questions related to the accuracy or integrity of any part of the work were appropriately investigated and resolved.

## Transparency Declaration

DAJ and ARM are the manuscript’s guarantors and they affirm that this is an honest, accurate, and transparent account of the study being reported and that no important aspects of the study have been omitted. All analyses were pre-specified in the study protocol.

## Data Sharing

the study dataset is available from.

