## Supplementary Materials for "Vitamin D supplementation to prevent acute respiratory infections: systematic review and meta-analysis of stratified aggregate data"

**Supplementary Material**

[13	Figure S3: Forest plot of RCTs comparing effects of vitamin D vs. placebo, reporting proportion of participants experiencing at least one acute respiratory infection, by baseline 25-hydroxyvitamin D level. A) <25.0 nmol/L; B) 25.0 to 49.9 nmol/L; C) 50.0 to 74.9 nmol/L, and D) ≥75.0 nmol/L.	18](#_Toc176339571)

[15	Figure S5: Forest plot of RCTs comparing effects of vitamin D vs. placebo, reporting proportion of participants experiencing at least one acute respiratory infection, by daily dose equivalents (<400 IU/day vs. 400-1000 IU/day vs. 1001-2000 IU/day vs. >2000 IU/day).	20](#_Toc176339573)

[17	Figure S7: Forest plot of RCTs comparing effects of vitamin D vs. placebo, reporting proportion of participants experiencing at least one acute respiratory infection, by the following age strata: A) <1 year; B) 1-15.99 years; C) 16-64.99 years, and D) ≥65 years	22](#_Toc176339575)

### 1 Search strategies

A. Medline

***Cochrane Highly Sensitive Search Strategy for identifying randomised controlled trials***

#1. randomized controlled trial [pt] OR controlled clinical trial [pt] OR randomized [tiab] OR placebo [tiab] OR drug therapy [sh] OR randomly [tiab] OR trial [tiab] OR groups [tiab]

#2. animals [mh] NOT humans [mh]

#3. #1 NOT #2

***Terms specific to vitamin D***

#4. Vitamin D OR vitamin D2 OR vitamin D3 OR cholecalciferol OR ergocalciferol OR

alphacalcidol OR alfacalcidol OR calcitriol OR paricalcitol OR doxerocalciferol

***Terms specific to acute respiratory infection***

#5. Acute Respiratory Infection OR Upper Respiratory Infection OR Lower Respiratory Infection OR Respiratory Tract Infection OR Common Cold OR Sinusitis OR Pharyngitis OR Laryngitis OR Laryngotracheobronchitis OR Tonsillitis OR peritonsillar abscess OR Croup OR Epiglottitis OR supraglottitis OR Otitis Media OR Pneumonia OR Bronchopneumonia OR Bronchitis OR Bronchiolitis OR Pleurisy OR Pleuritis OR Wheez* OR Respiratory OR Asthma

***Combination of terms to identify randomised controlled trials of vitamin D for the prevention of acute respiratory infection***

#3 AND #4 AND #5

B. EMBASE

***Terms for identifying randomised controlled trials***

#1. ‘randomized controlled trial’/exp OR ‘single blind procedure’/exp OR ‘double blind procedure’/exp OR ’crossover procedure’/exp

#2. random*:ab,ti OR placebo*:ab,ti OR crossover*:ab,ti OR ’cross over’:ab,ti OR allocat*:ab,ti OR ((singl* OR doubl*) NEXT/1 blind*):ab,ti OR trial:ti

#3. #1 OR #2

***Terms specific to vitamin D***

#4. *vitamin AND d OR vitamin AND d2 OR vitamin AND d3 OR cholecalciferol OR ergocalciferol OR alphacalcidol OR alfacalcidol OR calcitriol OR paricalcitol OR doxerocalciferol*

***Terms specific to acute respiratory infection***

#5. acute AND respiratory AND infection OR upper AND respiratory AND infection OR lower AND respiratory AND infection OR respiratory AND tract AND infection OR common AND cold OR sinusitis OR pharyngitis OR laryngitis OR laryngotracheobronchitis OR tonsillitis OR peritonsillar AND abscess OR croup OR epiglottitis OR supraglottitis OR otitis AND media OR pneumonia OR bronchopneumonia OR bronchitis OR bronchiolitis OR pleurisy OR pleuritis OR wheez* OR respiratory OR asthma

***Combination of terms to identify randomised controlled trials of vitamin D for the prevention of acute respiratory infection***

#3 AND #4 AND #5

C. Cochrane Central

***Terms specific to vitamin D***

#1. Vitamin D OR vitamin D2 OR vitamin D3 OR cholecalciferol OR ergocalciferol OR

alphacalcidol OR alfacalcidol OR calcitriol OR paricalcitol OR doxerocalciferol

***Terms specific to acute respiratory infection***

#2. Acute Respiratory Infection OR Upper Respiratory Infection OR Lower Respiratory Infection OR Respiratory Tract Infection OR Common Cold OR Sinusitis OR Pharyngitis OR Laryngitis OR Laryngotracheobronchitis OR Tonsillitis OR peritonsillar abscess OR Croup OR Epiglottitis OR supraglottitis OR Otitis Media OR Pneumonia OR Bronchopneumonia OR Bronchitis OR Bronchiolitis OR Pleurisy OR Pleuritis OR Wheez* OR Respiratory OR Asthma

***Combination of terms to identify randomised controlled trials of vitamin D for the prevention of acute respiratory infection***

#1 AND #2

D. Web of Science

TS =(Vitamin D OR vitamin D2 OR vitamin D3 OR cholecalciferol OR ergocalciferol OR alphacalcidol OR alfacalcidol OR calcitriol OR paricalcitol OR doxerocalciferol) AND TS =(Acute Respiratory Infection OR Upper Respiratory Infection OR Lower Respiratory Infection OR Respiratory Tract Infection OR Common Cold OR Sinusitis OR Pharyngitis OR Laryngitis OR Laryngotracheobronchitis OR Tonsillitis OR peritonsillar abscess OR Croup OR Epiglottitis OR supraglottitis OR Otitis Media OR Pneumonia OR Bronchopneumonia OR Bronchitis OR Bronchiolitis OR Pleurisy OR Pleuritis OR Wheez* OR Respiratory OR Asthma) AND TS =(placebo* or random* or clinical trial* or double blind* or single blind* or rct)

E. ClinicalTrials.gov

Vitamin D AND respiratory AND infection

### 2 Data Collection Processes

Summary data from trials which contributed to our previous meta-analysis of individual participant data,^1^ were extracted from our central database with permission from the Principal Investigators. Summary data relating to the primary outcome (overall and by sub-group) and secondary outcomes (overall only) from newly identified trials were requested from Principal Investigators. On receipt, they were assessed for consistency with associated publications. Study authors were contacted to provide missing data and to resolve any queries arising from these consistency checks. Once queries had been resolved, clean summary data were uploaded to the study database, which was held in STATA IC v14.2 (StataCorp, College Station, TX).

Data were extracted for the following variables: study setting, eligibility criteria, age, sex and baseline vitamin D status of participants, details of intervention and control regimens, trial duration, case definitions for ARI, and numbers of participants contributing data to statistical analyses. Follow-up summary data were requested for the proportions of participants experiencing one or more ARI during the trial, both overall and stratified by potential effect modifiers, where this was available. We also requested summary data on the proportions of participants who experienced one or more of the following events during the trial: upper respiratory infection (URI); lower respiratory infection (LRI); Emergency Department attendance and/or hospital admission for ARI; death due to ARI or respiratory failure; use of antibiotics to treat an ARI; absence from work or school due to ARI; a serious adverse event; death due to any cause; and potential adverse reactions to vitamin D (hypercalcaemia and renal stones).

### 3 Sources of support for participating trials

The trial by Aglipay and colleagues was supported by the competitive grants from the Canadian Institutes of Health Research Institutes of Human Development, Child and Youth Health and Nutrition, Metabolism and Diabetes (grant number MOP-114945) and the Thrasher Research Fund (award number 9113).

The trial by Aloia and colleagues was supported by the National Institute of Aging (grant number R01-AG032440-01A2).

The trial by Arihiro and colleagues was supported by the Ministry of Education, Culture, Sports, Science, and Technology in the Japan-Supported Program for the Strategic Research Foundation at Private Universities and funding from the Department of Gastroenterology and Hepatology, Jikei University of Medicine, Tokyo, Japan.

The trial by Bergman and colleagues was supported by grants from the Swedish Research Council, the Strategic Research Foundation (SSF), the Swedish Heart and Lung foundation, Karolinska Institutet, Stockholm County Council and the Swedish Cancer Society as well as by the Magnus Bergwall and Åke Wiberg foundations.

The trial by Bischoff-Ferrari and colleagues was supported by grants from the Seventh Framework Program of the European Commission (grant agreement 278588), the University of Zurich (Chair for Geriatric Medicine and Aging Research), DSM Nutritional Products, Roche, NESTEC, Pfizer, and Streuli.

The trials by Camargo and colleagues were supported by a grant for the Blue Sky Study from an anonymous foundation and the Massachusetts General Hospital; and the Health Research Council of New Zealand (grant number 10/400) and the Accident Compensation Corporation of New Zealand. The trial by Camargo, Manson, and colleagues was supported by National Institutes of Health (NIH) (grant numbers U01 CA138962, R01 CA138962, R01 AT011729, R01 AI093723, K24 HL136852). Pharmavite, Pronova BioPharma/BASF, and Quest Diagnostics provided study medications and diagnostic services.

The trial by Ganmaa and colleagues was supported by the National Institutes of Health (Grant Number 1R01HL122624-01).

The trial by Ginde and colleagues was supported by NIH/NIA grant K23AG040708, NIH/NCATS Colorado CTSA Grant UL1TR001082, and the American Geriatrics Society Jahnigen Career Development Scholars Award.

The trial by Golan-Tripto and colleagues was supported by the Soroka JNF UK Clinical Research Scholar Program.

The trial by Goodall and colleagues was supported in part by the Canadian Institutes of Health Research [OPP 86940] and with in-kind support from Copan Italia, Bresica Italy.

The trial by Grant and colleagues was supported by the Health Research Council of New Zealand, Grant Number 09/215R.

The trial by Gupta and colleagues was supported by the Indian Council of Medical Research, New Delhi.

The trial by Hauger and colleagues was supported by Lundbeckfonden (grant number R180-2014-3481), by Brødrene Hartmann’s Fund (A26842), and by the European Commission (FP7/2007–2013) under Grant Agreement 613977 for the ODIN Integrated Project (Food-based solutions for optimal vitamin D nutrition and health through the life cycle).

The trial by Hibbs and colleagues was supported by the National Heart, Lung, and Blood Institute and Office of Dietary Supplements (grant number R01HL109293).

The trial by Lee and colleagues was supported by the US Food and Drug Administration Orphan Product Development (grant number R01FD003894).

The trial by Loeb and colleagues was supported by the Institute for Infectious Diseases Research at McMaster University.

The trials by Manaseki-Holland and colleagues were supported by New Zealand Aid (ref: GRA/470/2) and The Wellcome Trust (ref: 082476).

The trial by Mandlik and colleagues was supported by a core grant from the Hirabai Cowasji Jehangir Medical Research Institute.

The trials by Martineau and colleagues were supported by the National Institute for Health Research under its Programme Grants for Applied Research Programme (Reference Number RP-PG-0407-10398).

The trial by Murdoch and colleagues was supported by the Health Research Council of New Zealand, grant number 09/302.

The trial by Pham and colleagues was supported by project grants from the National Health and Medical Research Council (grant numbers GNT1046681 and GNT1120682).

The trial by Reyes and colleagues was supported by Fondo Nacional de Investigacion y Desarrollo en Salud (ref. SA13I20173).

The trial by Rosendahl and colleagues was supported by the Foundation for Pediatric Research, the Finnish Medical Foundation, Governmental Subsidy for Clinical Research, the Päivikki and Sakari Sohlberg Foundation, the Academy of Finland, the Sigrid Jusélius Foundation, the Folkhälsan Research Foundation, the Novo Nordisk Foundation, the Orion Research Foundation, and Barncancerfonden.

The trial by Rake and colleagues was supported by the National Institute for Health Research Health Technology Assessment programme (ref. HTA 08/116/48).

The trial by Rees and colleagues was supported by the National Cancer Institute at the National Institutes of Health (grant numbers CA098286 and CA098286-S).

The trial by Shimizu and colleagues was supported by the FANCL Corporation.

The trial by Simpson and colleagues was supported by the Royal Hobart Hospital Research Foundation.

The trial by Tachimoto and colleagues was supported by the Ministry of Education, Culture, Sports, Science, and Technology in the Japan-Supported Program for the Strategic Research Foundation at Private Universities, JSPH KAKENHI Grant Number 23591553, and funding from Jikei University of Medicine, Tokyo, Japan.

The trial by Tran and colleagues was supported by the National Health and Medical Research Council of Australia, grant 613655.

The trial by Trilok Kumar and colleagues was supported by the Department of Biotechnology, Government of India (ref BT/PR-PR7489/PID/20/285/2006), Nutrition Third World and Sight and Life

### 4 Registration and Ethical Approvals

Research Ethics Committee approval to conduct this meta-analysis was not required in the UK; local ethical permission to contribute data from primary trials was required and obtained for studies by Camargo *et al,*^2^ (The Ethics Review Committee of the Mongolian Ministry of Health), Murdoch *et al,*^3^ (Southern Health and Disability Ethics Committee, ref. URB/09/10/050/AM02), Rees *et al,*^4^ (Committee for the Protection of Human Subjects, Dartmouth College, USA; Protocol # 24381), Tachimoto *et al,*^5^ (Ethics committee of the Jikei University School of Medicine, ref 26-333: 7839), Tran *et al,*^6^ (QIMR Berghofer Medical Research Institute Human Research Ethics Committee, P1570) and Urashima *et al,*^7^^,^^8^ (Ethics committee of the Jikei University School of Medicine, ref 26-333: 7839).

### 5 Exploration of variation in effects: meta-regression

To investigate factors associated with heterogeneity of effect between statistically significant subgroups of trials, we performed multivariable meta-regression analysis on trial-level characteristics, namely, dose frequency, dose size, trial duration and age at enrolment, to produce an adjusted odds ratio, a 95% CI and a P value for interaction for each factor. Independent variables were dichotomised to create a more parsimonious model (baseline serum 25(OH)D of <25 vs. ≥25 nmol/L; administration of daily vs. non-daily doses; daily equivalent of ≤1000 IU vs. >1000 IU; trial duration of ≤12 vs. >12 months, and participant age of <16.00 vs. ≥16.00 years at enrolment). The meta-regression analysis excluded data from two placebo-controlled trials that included higher-dose, lower-dose and placebo arms,^6^^,^^9^ (since the higher-dose and lower-dose arms in these studies spanned the 1,000 IU/day cut-off), and four placebo-controlled trials that enrolled participants aged below and above the age cut-off of 16 years.^7^^,^^10-12^ These factors rendered these trials unclassifiable for the purposes of the meta-regression analysis.

### 6 Table S1: Reasons for exclusion of potentially relevant studies

| **First author, year or clinicaltrials.gov registration number** | **Reason for exclusion** |
| --- | --- |
| Somnath, 2017^13^ | Ineligible: open-label trial |
| Jung, 2018^14^ | Ineligible: ARI outcome not pre-specified |
| Ramos-Martínez, 2018^15^ | Ineligible: intervention was administration of 1,25-dihydroxyvitamin D |
| Zhou, 2018^16^ | Ineligible: open-label trial |
| Hueniken, 2019^17^ | Ineligible: same trial as Aglipay et al^18^ |
| Singh, 2019^19^ | Ineligible: open-label trial |
| Jolliffe, 2022^20^ | Ineligible: open-label trial |
| van Helmond^21^ | Ineligible: open-label trial |
| NCT05037253 | Ineligible: open-label trial |
| NCT04810949 | Ineligible: open-label trial |
| Villasis-Keever^22^ | Eligible: unresponsive to invitation to contribute data |
| NCT04386850 | Potentially eligible: status of completion could not be determined |
| NCT03956732 | Ineligible: second intervention administered to vitamin D arm |

### 7 Table S2: Risk of Bias Assessment

|  | **Sequence generation** | **Allocation concealment** | **Blinding of participants and personnel** | **Blinding of outcome assessment** | **Incomplete outcome data** | **Selective reporting** | **Other bias** |
| --- | --- | --- | --- | --- | --- | --- | --- |
| Li-Ng 2009^23^ | **✓** | **✓** | **✓** | **✓** | **✓** | **✓** | **✓** |
| Urashima 2010^8^ | **✓** | **✓** | **✓** | **✓** | **✓** | **✓** | **✓** |
| Manaseki-Holland 2010^24^ | **✓** | **✓** | **✓** | **✓** | **✓** | **✓** | **✓** |
| Laaksi 2010^25^ | **✓** | **✓** | **✓** | **✓** | **?** | **✓** | **✓** |
| Majak 2011^10^ | **✓** | **✓** | **✓** | **✓** | **✓** | **✓** | **✓** |
| Trilok-Kumar 2011^26^ | **✓** | **✓** | **✓** | **✓** | **✓** | **✓** | **✓** |
| Lehouck 2012^27^ | **✓** | **✓** | **✓** | **✓** | **✓** | **✓** | **✓** |
| Manaseki-Holland 2012^28^ | **✓** | **✓** | **✓** | **✓** | **✓** | **✓** | **✓** |
| Camargo 2012^2^ | **✓** | **✓** | **✓** | **✓** | **✓** | **✓** | **✓** |
| Murdoch 2012^3^ | **✓** | **✓** | **✓** | **✓** | **✓** | **✓** | **✓** |
| Bergman 2012^29^ | **✓** | **✓** | **✓** | **✓** | **✓** | **✓** | **✓** |
| Marchisio 2013^30^ | **✓** | **✓** | **✓** | **✓** | **✓** | **✓** | **✓** |
| Rees 2013^4^ | **✓** | **✓** | **✓** | **✓** | **✓** | **✓** | **✓** |
| Tran 2014^6^ | **✓** | **✓** | **✓** | **✓** | **✓** | **✓** | **✓** |
| Goodall 2014^31^ | **✓** | **✓** | **✓** | **✓** | **✓** | **✓** | **✓** |
| Urashima 2014^7^ | **✓** | **✓** | **✓** | **✓** | **✓** | **✓** | **✓** |
| Grant 2014^32^ | **✓** | **✓** | **✓** | **✓** | **✓** | **✓** | **✓** |
| Martineau 2015a^33^ [ViDiCO] | **✓** | **✓** | **✓** | **✓** | **✓** | **✓** | **✓** |
| Martineau 2015b^34^ [ViDiAs] | **✓** | **✓** | **✓** | **✓** | **✓** | **✓** | **✓** |
| Martineau 2015c^35^ [ViDiFlu] | **✓** | **✓** | **✓** | **✓** | **✓** | **✓** | **✓** |
| Simpson 2015^36^ | **✓** | **✓** | **✓** | **✓** | **✓** | **✓** | **✓** |
| Dubnov-Raz 2015^11^ | **✓** | **✓** | **✓** | **✓** | **?** | **✓** | **✓** |
| Denlinger 2016^37^ | **✓** | **✓** | **✓** | **✓** | **✓** | **✓** | **✓** |
| Tachimoto 2016^5^ | **✓** | **✓** | **✓** | **✓** | **✓** | **✓** | **✓** |
| Ginde 2016^38^ | **✓** | **✓** | **✓** | **✓** | **✓** | **✓** | **✓** |
| Gupta 2016^39^ | **✓** | **✓** | **✓** | **✓** | **✓** | **✓** | **✓** |
| Aglipay 2017 ^18^ | **✓** | **✓** | **✓** | **✓** | **✓** | **✓** | **✓** |
| Arihiro 2018 ^40^ | **✓** | **✓** | **✓** | **✓** | **✓** | **✓** | **✓** |
| Hibbs 2018 ^41^ | **✓** | **✓** | **✓** | **✓** | **✓** | **✓** | **✓** |
| Lee 2018 ^42^ | **✓** | **✓** | **✓** | **✓** | **✓** | **✓** | **✓** |
| Loeb 2018 ^12^ | **✓** | **✓** | **✓** | **✓** | **✓** | **✓** | **✓** |
| Rosendahl 2018 ^43^ | **✓** | **✓** | **✓** | **✓** | **✓** | **✓** | **✓** |
| Shimizu 2018 ^44^ | **✓** | **✓** | **✓** | **✓** | **✓** | **✓** | **✓** |
| Aloia 2019 ^45^ | **✓** | **✓** | **✓** | **✓** | **✓** | **✓** | **✓** |
| Hauger 2019 ^46^ | **✓** | **✓** | **✓** | **✓** | **✓** | **✓** | **✓** |
| Bischoff-Ferrari^47^ | **✓** | **✓** | **✓** | **✓** | **✓** | **✓** | **✓** |
| Camargo 2020 ^48^ | **✓** | **✓** | **✓** | **✓** | **✓** | **✓** | **✓** |
| Ganmaa 2020^49^ | **✓** | **✓** | **✓** | **✓** | **✓** | **✓** | **✓** |
| Mandlik 2020 ^50^ | **✓** | **✓** | **✓** | **✓** | **✓** | **✓** | **✓** |
| Pham 2020^51^ | **✓** | **✓** | **✓** | **✓** | **✓** | **✓** | **✓** |
| Rake 2020 ^52^ | **✓** | **✓** | **✓** | **✓** | **✓** | **✓** | **✓** |
| Golan‐Tripto (unpublished) ^53^ | **✓** | **✓** | **✓** | **✓** | **?** | **N/A** | **✓** |
| Reyes (unpublished) ^9^ | **✓** | **✓** | **✓** | **✓** | **?** | **N/A** | **✓** |
| Huang 2022 | **✓** | **✓** | **✓** | **?** | **?** | **?** | **✓** |
| Manson 2023 | **✓** | **✓** | **✓** | **✓** | **✓** | **✓** | **✓** |
| Ducharme (unpublished) | **✓** | **✓** | **✓** | **✓** | **✓** | **N/A** | **✓** |

**✓ = low risk of bias; ? = unclear risk of bias, N/A = not applicable (unpublished)**

### 8 Table S3: Multivariable meta-regression model for proportion of participants experiencing at least one acute respiratory tract infection, by trial-level subgroups.

| **Variables** | **No of trials^[a]^** | **Proportion with ≥1 ARI, intervention group (%)** | **Proportion with ≥1 ARI, control group (%)** | **Odds ratio (95% CI)^[b]^** | **I^2^ %** | **P value for heterogeneity** | **Adjusted odds ratio (95% CI)^[c]^** | **P value for interaction^[c]^** |
| --- | --- | --- | --- | --- | --- | --- | --- | --- |
| **Dosing frequency** | | | | | | | | |
| Daily | 21 | 2572/10920 (23.6) | 2569/10632 (24.2) | 0.84 (0.73 to 0.97) | 44.8 | 0.01 | 1.01 (0.87 to 1.17) | 0.91 |
| Not daily | 18 | 12445/19775 (62.9) | 12452/19668 (63.3) | 0.98 (0.93 to 1.02) | 0.0 | 0.56 | Referent |  |
| **Daily dose equivalent, IU** | | | | | | | | |
| ≤1000 | 12 | 1107/2310 (47.9) | 1100/2128 (51.7) | 0.74 (0.60 to 0.90) | 35.3 | 0.11 | 0.89 (0.73 to 1.08) | 0.23 |
| >1000 | 26 | 13785/28252 (48.8) | 13862/28111 (49.3) | 0.98 (0.93 to 1.04) | 14.7 | 0.25 | Referent |  |
| **Trial duration, months** | | | | | | | | |
| ≤12 | 31 | 2847/12615 (22.6) | 2766/12063 (23.0) | 0.85 (0.76 to 0.95) | 32.7 | 0.04 | Referent | 0.20 |
| >12 | 8 | 12355/18477 (66.9) | 12351/18434 (67.0) | 0.99 (0.95 to 1.04) | 0.0 | 0.95 | 1.10 (0.95 to 1.28) |  |
| **Age, years** | | | | | | | | |
| ≥16 | 21 | 9845/21674 (45.4) | 9914/21545 (46.0) | 0.96 (0.90 to 1.03) | 17.6 | 0.23 | 0.99 (0.86 to 1.14) | 0.86 |
| <16 | 14 | 5076/8258 (61.5) | 5026/8045 (62.5) | 0.94 (0.88 to 1.00) | 38.5 | 0.07 | Referent |  |

[a] Data from two trials that included higher-dose, lower-dose and placebo arms^6^^,^^9^ are excluded, since the higher-dose and lower-dose arms in these studies spanned the 1,000 IU/day cut-off, and data from four placebo-controlled trials ^7^^,^^10-12^were excluded, since some participants spanned the 16 years of age cut-off, rendering all these trials unclassifiable for the purposes of this analysis.

[b] Within-sub-group odds ratios from random effects model adjusting for study weights.

[c] adjusted odds ratios and P values for interaction from multivariable meta-regression model including dichotomised variables for dose frequency, size and trial duration.

### 9 Table S4: Summary of Findings Table

|  | | | | | |
| --- | --- | --- | --- | --- | --- |
| **Vitamin D_3_ compared to placebo for prevention of acute respiratory infection (ARI)** | | | | | |
| **Population**: children and adults of any age, sex or ethnic origin, with or without co-morbidity  **Setting**: Eighteen countries on four continents (Asia, Australasia, Europe, North America)  **Intervention**: oral vitamin D_3_ (cholecalciferol) supplementation  **Comparison**: oral placebo | | | | | |
| Outcomes | **Anticipated absolute effects^*^** (95% CI) | | Relative effect (95% CI) | № of participants  (studies) | Quality of the evidence (GRADE) |
|  | **Risk with placebo** | **Risk with Vitamin D** |  |  |  |
| Proportion with at least one ARI, all participants | 496 per 1,000 | 480 per 1,000 (464 to 496) | OR 0.94 (0.88 to 1.00) | 61589 (40 RCTs) | ⨁⨁⨁ MODERATE |
| Proportion with at least one ARI, participants in placebo-controlled trials with duration ≤12 months investigating daily dosing with 400-1,000 IU vitamin D_3_/day | 572 per 1,000 | 436 per 1,000 (375 to 500) | OR 0.58 (0.45 to 0.75) | 1232 (8 RCTs) | ⨁⨁⨁ MODERATE |
| Proportion with at least one hospital admission or emergency department attendance due to ARI | 14 per 1,000 | 12 per 1,000 (10 to 16) | OR 0.90 (0.71 to 1.14) | 21846  (20 RCTs) | ⨁⨁⨁ MODERATE |
| Proportion with serious adverse event, any cause | 73 per 1,000 | 70 per 1,000 (66 to 75) | OR 0.96 (0.90 to 1.04) | 45181 (38 RCTs) | ⨁⨁⨁ MODERATE |
| Proportion of deaths due to ARI or respiratory failure | 1 per 1,000 | 1 per 1,000 (0 to 1) | OR 1.03 (0.61 to 1.75) | 28860 (35 RCTs) | ⨁⨁⨁ MODERATE |
| ***The risk in the intervention group** (and its 95% confidence interval) is based on the assumed risk in the comparison group and the **relative effect** of the intervention (and its 95% CI).   **CI:** Confidence interval; **OR:** Odds ratio | | | | | |
| **GRADE Working Group grades of evidence** **High quality:** We are very confident that the true effect lies close to that of the estimate of the effect **Moderate quality:** We are moderately confident in the effect estimate: The true effect is likely to be close to the estimate of the effect, but there is a possibility that it is substantially different **Low quality:** Our confidence in the effect estimate is limited: The true effect may be substantially different from the estimate of the effect **Very low quality:** We have very little confidence in the effect estimate: The true effect is likely to be substantially different from the estimate of effect | | | | | |

### 10 Table S5: Results of exploratory sensitivity analyses excluding placebo-controlled trials at unclear risk of bias and studies investigating ARI incidence as a secondary outcome.

| **Reason for exclusion (number of excluded studies)** | **No. of studies included** | **Proportion with ≥1 ARI, intervention group (%)** | **Proportion with ≥1 ARI, control group (%)** | **Odds ratio (95% CI)** | **I^2^ %** | **P value for heterogeneity** |
| --- | --- | --- | --- | --- | --- | --- |
| Studies at unclear risk of bias (n=4) | 36 | 15027/30730 (48.9) | 14989/30228 (49.6) | 0.95 (0.90 to 1.01) | 22.3 | 0.12 |
| Studies with ARI incidence as secondary outcome (n=19) | 21 | 1804/5080 (35.5) | 1767/4895 (36.1) | 0.90 (0.79 to 1.02) | 18.2 | 0.22 |

### 11 Figure S1: Cates plot illustrating reduction in risk of one or more acute respiratory infections with vitamin D supplementation vs. placebo, overall (primary comparison).

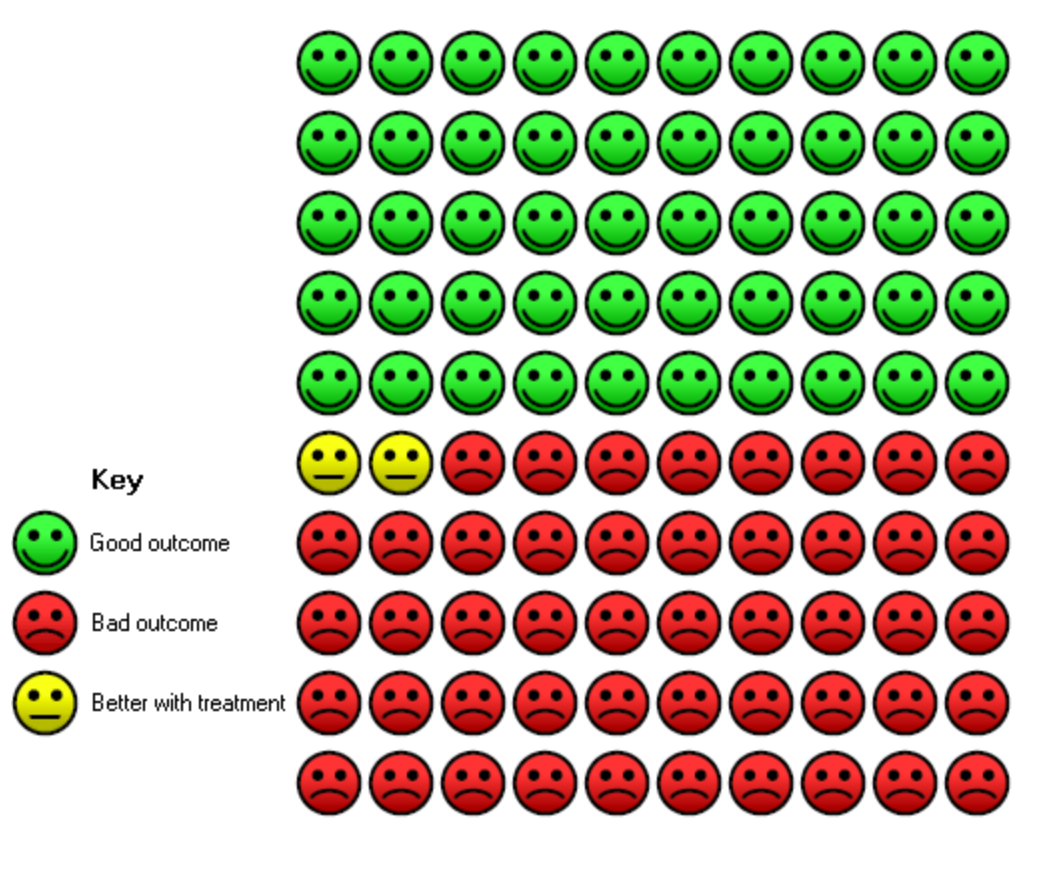

### 12 Figure S2: Forest plot of RCTs comparing effects of higher- vs. lower-dose vitamin D, reporting proportion of participants experiencing at least one acute respiratory infection.

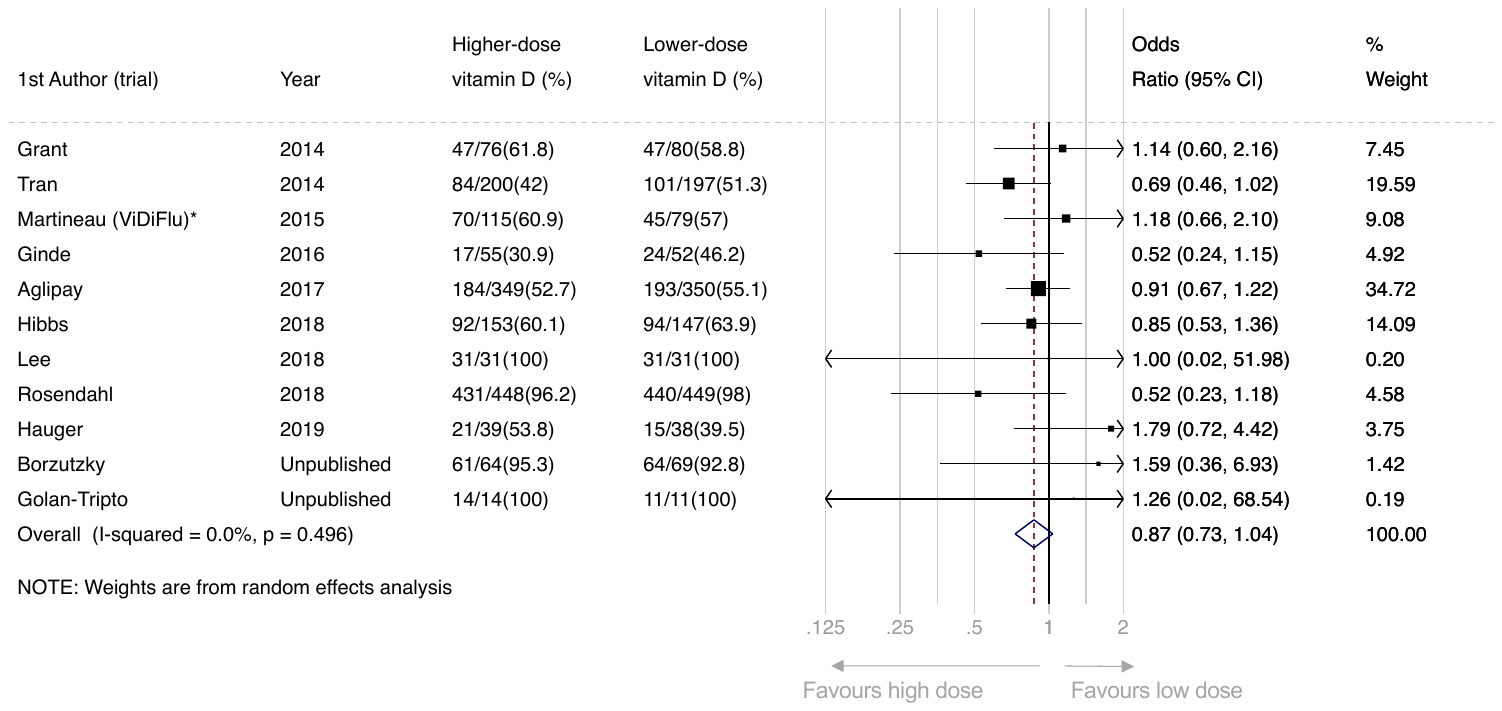

*This analysis includes data from the subset of ViDiFlu trial participants who were randomised to higher- vs. lower-dose vitamin D.

### 13 Figure S3: Forest plot of RCTs comparing effects of vitamin D vs. placebo, reporting proportion of participants experiencing at least one acute respiratory infection, by baseline 25-hydroxyvitamin D level. A) <25.0 nmol/L; B) 25.0 to 49.9 nmol/L; C) 50.0 to 74.9 nmol/L, and D) ≥75.0 nmol/L.

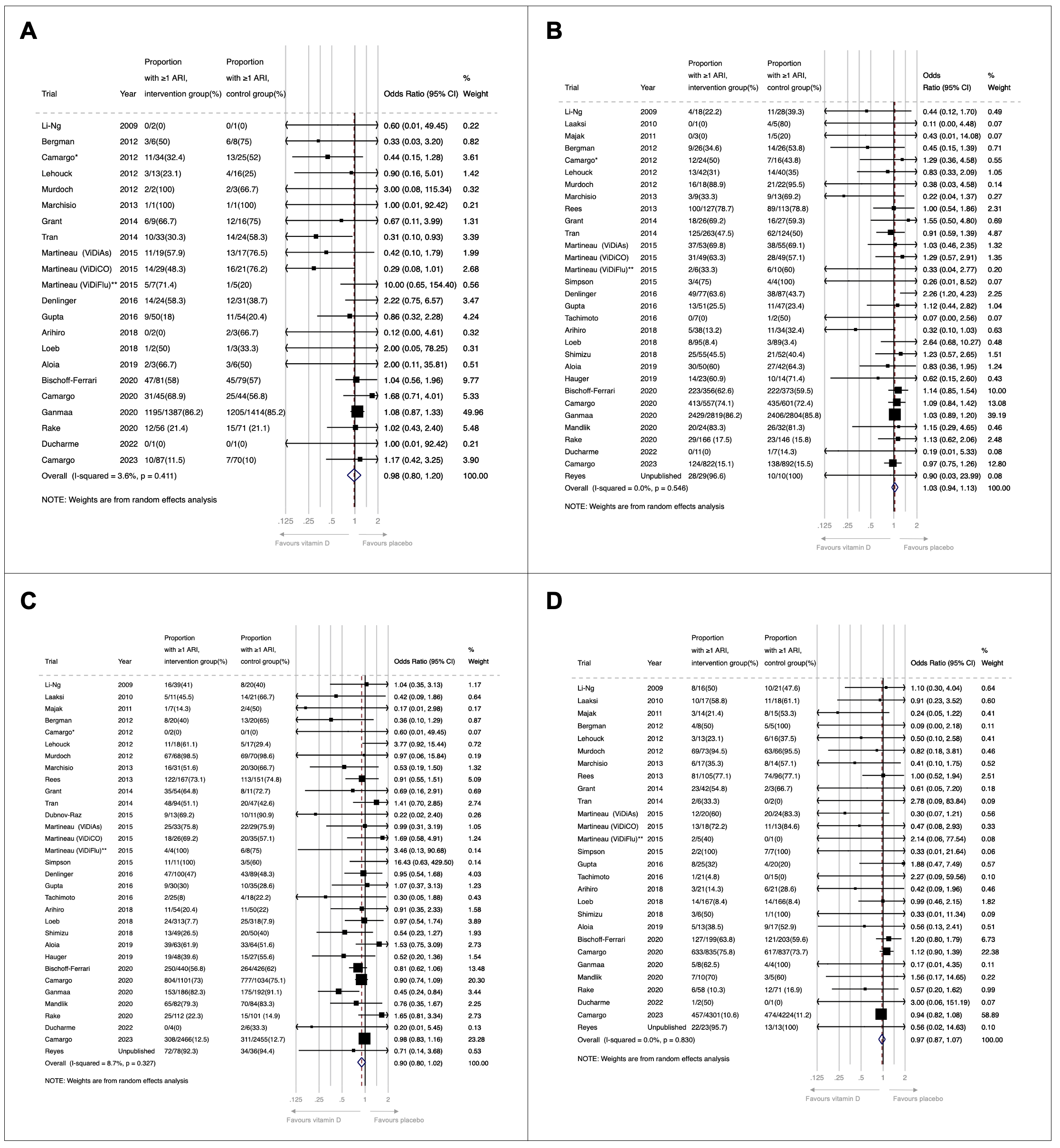

*Proportions for this trial were corrected for cluster randomisation using the calculated design effect of 4.12 for participants and clusters in the <25.0 nmol/L group (A) and the design effect of 1.23 for participants and clusters in the 25.0 to 49.9 nmol/L group (B). Design effect was incalculable for the 50.0 to 74.9 nmol/L group (C) due to the lack of power. **This analysis includes data from the subset of ViDiFlu trial participants who were randomised to vitamin D vs. placebo control; correction for cluster randomisation was not possible due to the lack of power.

### 14 Figure S4: Forest plot of RCTs comparing effects of vitamin D vs. placebo, reporting proportion of participants experiencing at least one acute respiratory infection, by frequency of supplementation (daily vs. weekly vs. monthly to 3-monthly)

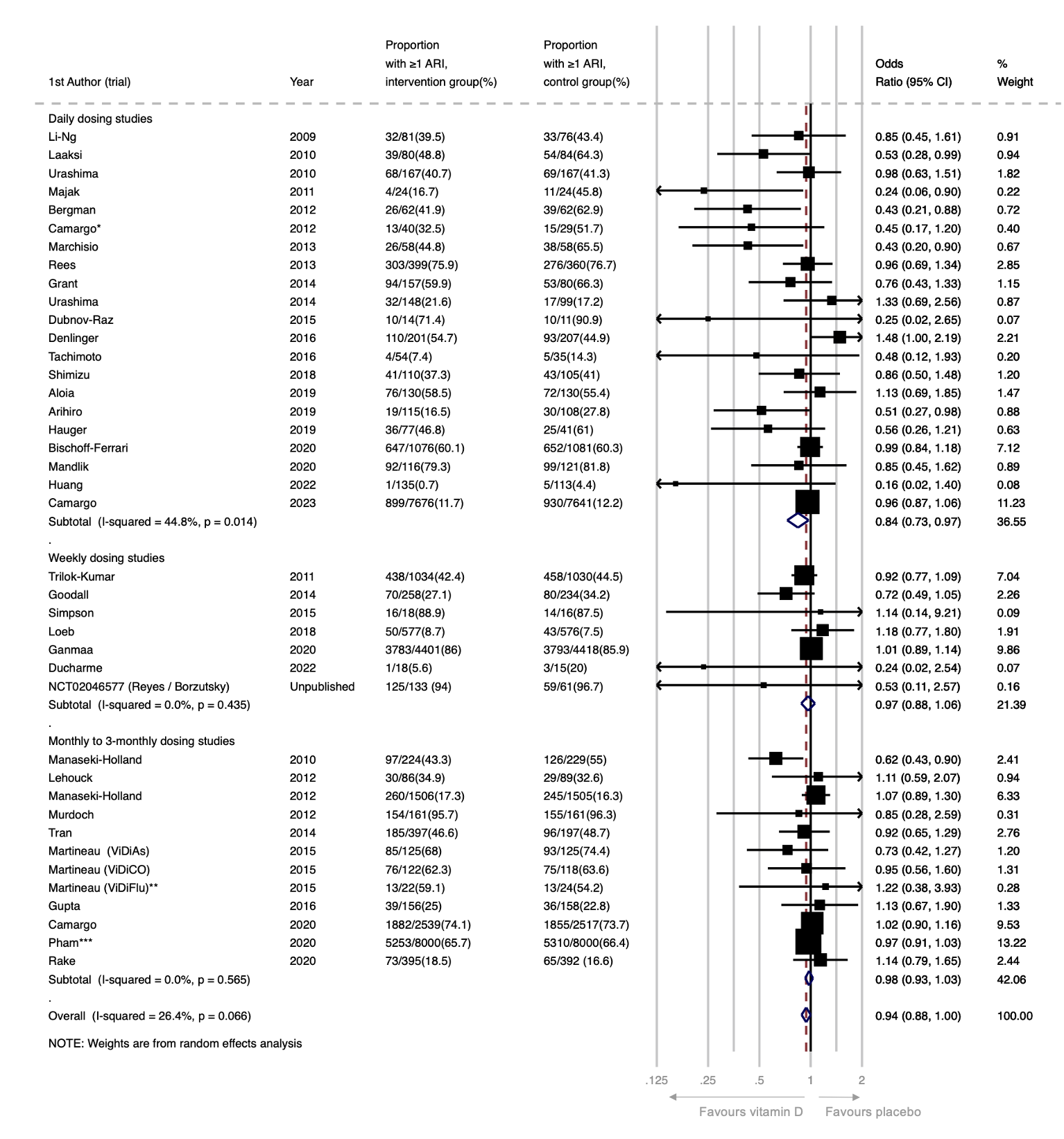

*Proportions for this trial were corrected for cluster randomisation using the calculated design effect of 3.49. **This analysis includes data from the subset of ViDiFlu trial participants who were randomised to vitamin D vs. placebo control; correction for cluster randomisation was not possible due to the lack of power. ***For this trial, participants were asked to report the occurrence of ARTI during the one month prior to completing each annual survey (max surveys=5). The numerator is the number of people who reported an ARTI on at least one survey. The ARTI outcomes for people who completed fewer than 5 surveys and who did not report an ARTI (N=2239; 14%) were estimated based on the % affected among those who completed all 5 surveys (N=12,152; 76%).

### 15 Figure S5: Forest plot of RCTs comparing effects of vitamin D vs. placebo, reporting proportion of participants experiencing at least one acute respiratory infection, by daily dose equivalents (<400 IU/day vs. 400-1000 IU/day vs. 1001-2000 IU/day vs. >2000 IU/day).

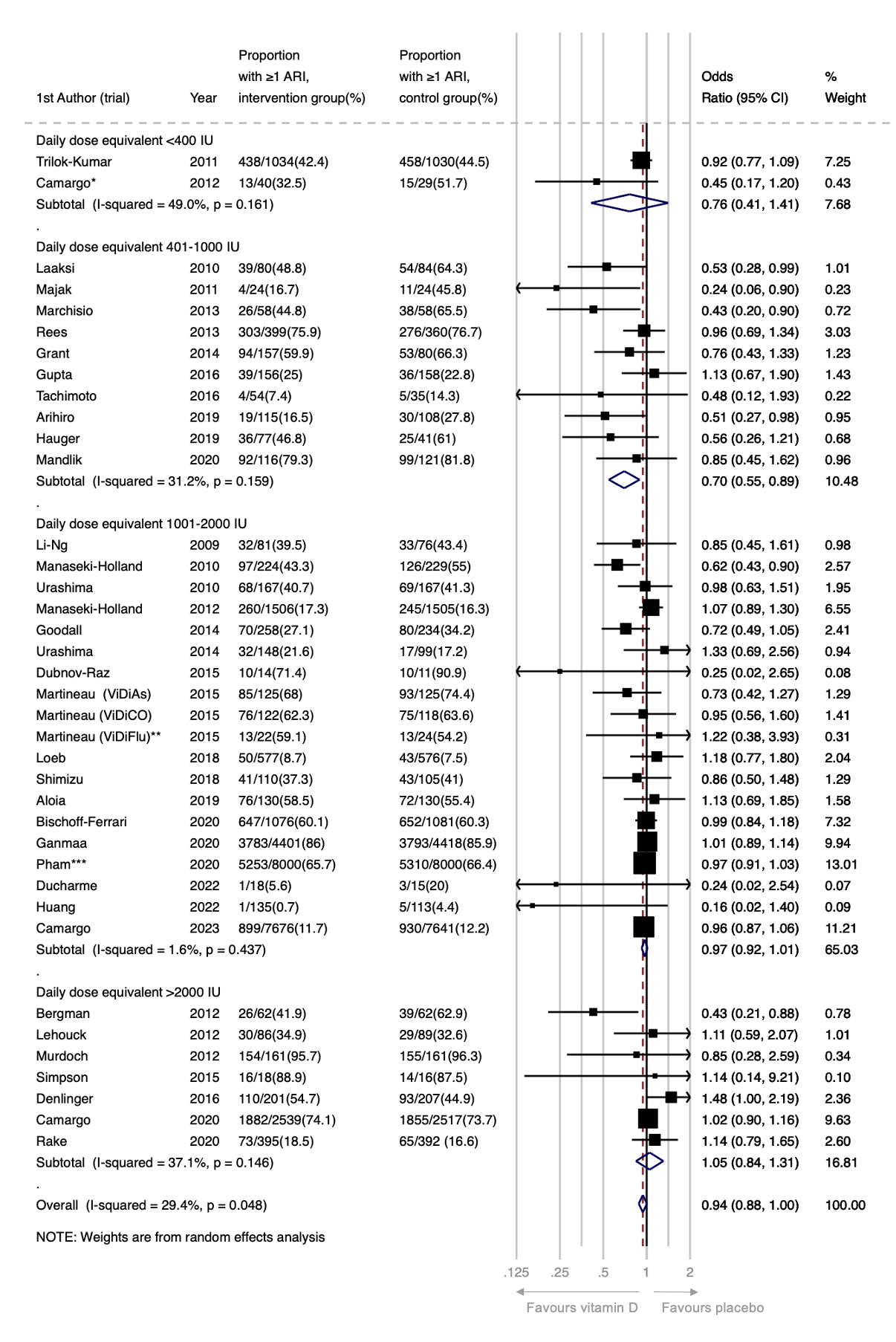

*Proportions for this trial were corrected for cluster randomisation using the calculated design effect of 3.49. **This analysis includes data from the subset of ViDiFlu trial participants who were randomised to vitamin D vs. placebo control; correction for cluster randomisation was not possible due to the lack of power. ***For this trial, participants were asked to report the occurrence of ARTI during the one month prior to completing each annual survey (max surveys=5). The numerator is the number of people who reported an ARTI on at least one survey. The ARTI outcomes for people who completed fewer than 5 surveys and who did not report an ARTI (N=2239; 14%) were estimated based on the % affected among those who completed all 5 surveys (N=12,152; 76%).

### 16 Figure S6: Forest plot of RCTs comparing effects of vitamin D vs. placebo, reporting proportion of participants experiencing at least one acute respiratory infection, by trial duration (≤12 months vs. >12 months).

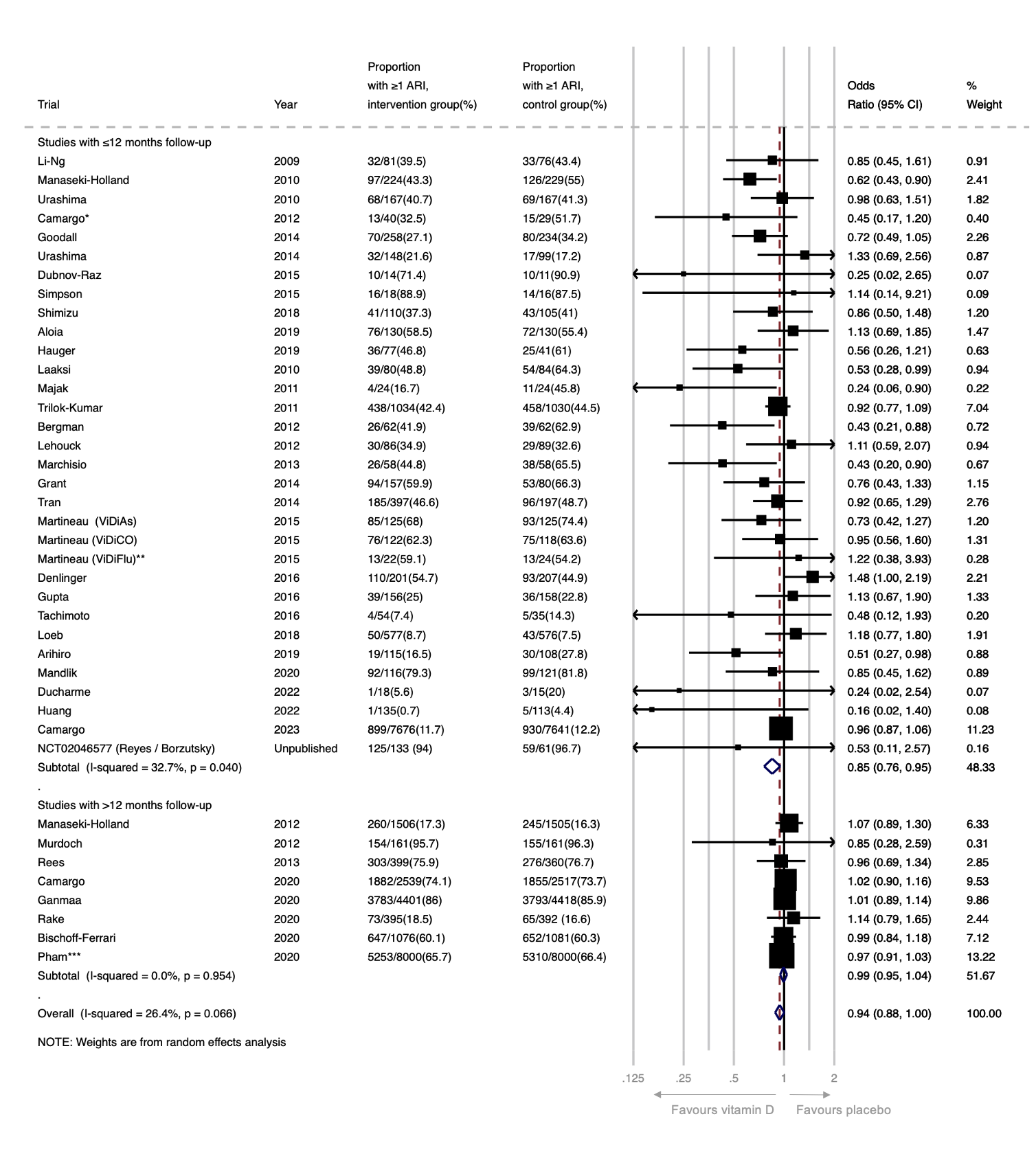

*Proportions for this trial were corrected for cluster randomisation using the calculated design effect of 3.49. **This analysis includes data from the subset of ViDiFlu trial participants who were randomised to vitamin D vs. placebo control; correction for cluster randomisation was not possible due to the lack of power. ***For this trial, participants were asked to report the occurrence of ARTI during the one month prior to completing each annual survey (max surveys=5). The numerator is the number of people who reported an ARTI on at least one survey. The ARTI outcomes for people who completed fewer than 5 surveys and who did not report an ARTI (N=2239; 14%) were estimated based on the % affected among those who completed all 5 surveys (N=12,152; 76%).

### 17 Figure S7: Forest plot of RCTs comparing effects of vitamin D vs. placebo, reporting proportion of participants experiencing at least one acute respiratory infection, by the following age strata: A) <1 year; B) 1-15.99 years; C) 16-64.99 years, and D) ≥65 years

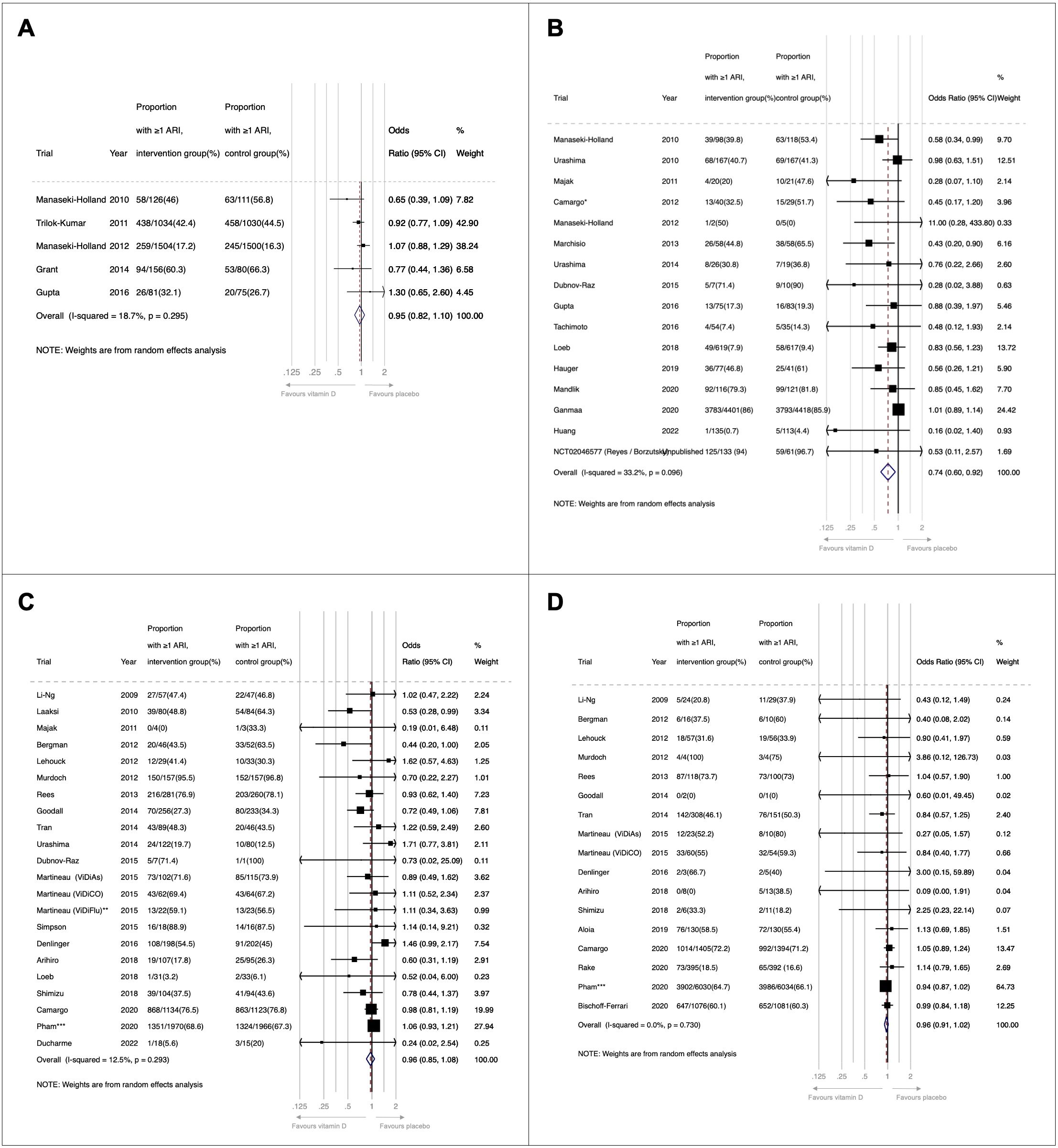

*Proportions for this trial were corrected for cluster randomisation using the calculated design effect of 3.49. **This analysis includes data from the subset of ViDiFlu trial participants who were randomised to vitamin D vs. placebo control. ***For this trial, participants were asked to report the occurrence of ARTI during the one month prior to completing each annual survey (max surveys=5). The numerator is the number of people who reported an ARTI on at least one survey. The ARTI outcomes for people who completed fewer than 5 surveys and who did not report an ARTI (N=2239; 14%) were estimated based on the % affected among those who completed all 5 surveys (N=12,152; 76%). Age-stratified data for Camargo et al. 2023 were not available.

### 18 Figure S8: Forest plot of RCTs comparing effects of vitamin D vs. placebo, reporting proportion of participants experiencing at least one acute respiratory infection, by presence or absence of airway disease comorbidity.

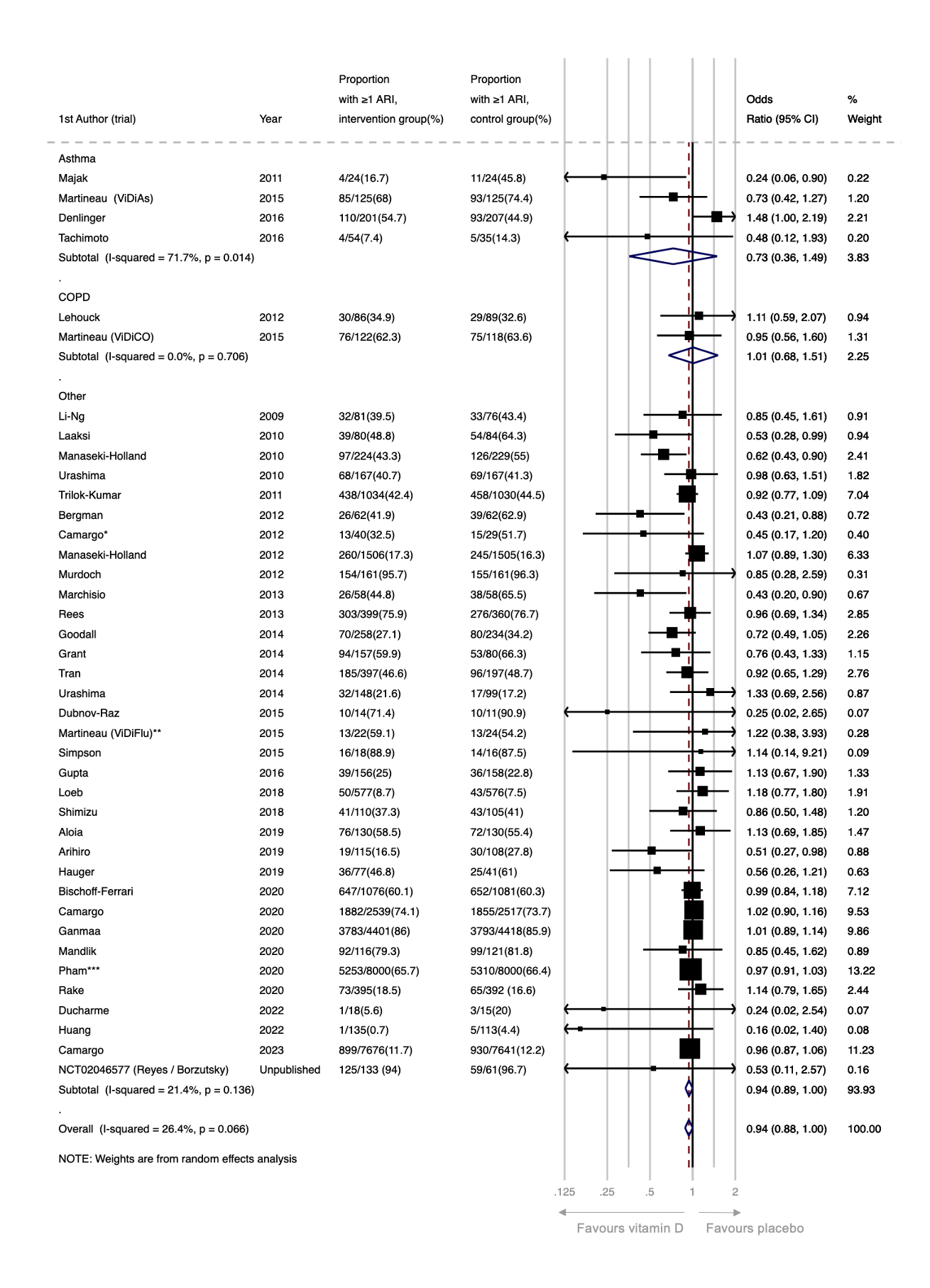

*Proportions for this trial were corrected for cluster randomisation using the calculated design effect of 3.49. **This analysis includes data from the subset of ViDiFlu trial participants who were randomised to vitamin D vs. placebo control; correction for cluster randomisation was not possible due to the lack of power. ***For this trial, participants were asked to report the occurrence of ARTI during the one month prior to completing each annual survey (max surveys=5). The numerator is the number of people who reported an ARTI on at least one survey. The ARTI outcomes for people who completed fewer than 5 surveys and who did not report an ARTI (N=2239; 14%) were estimated based on the % affected among those who completed all 5 surveys (N=12,152; 76%).

### 19 Figure S9: Funnel plot of placebo-controlled RCTs reporting proportion of participants experiencing 1 or more acute respiratory infection.

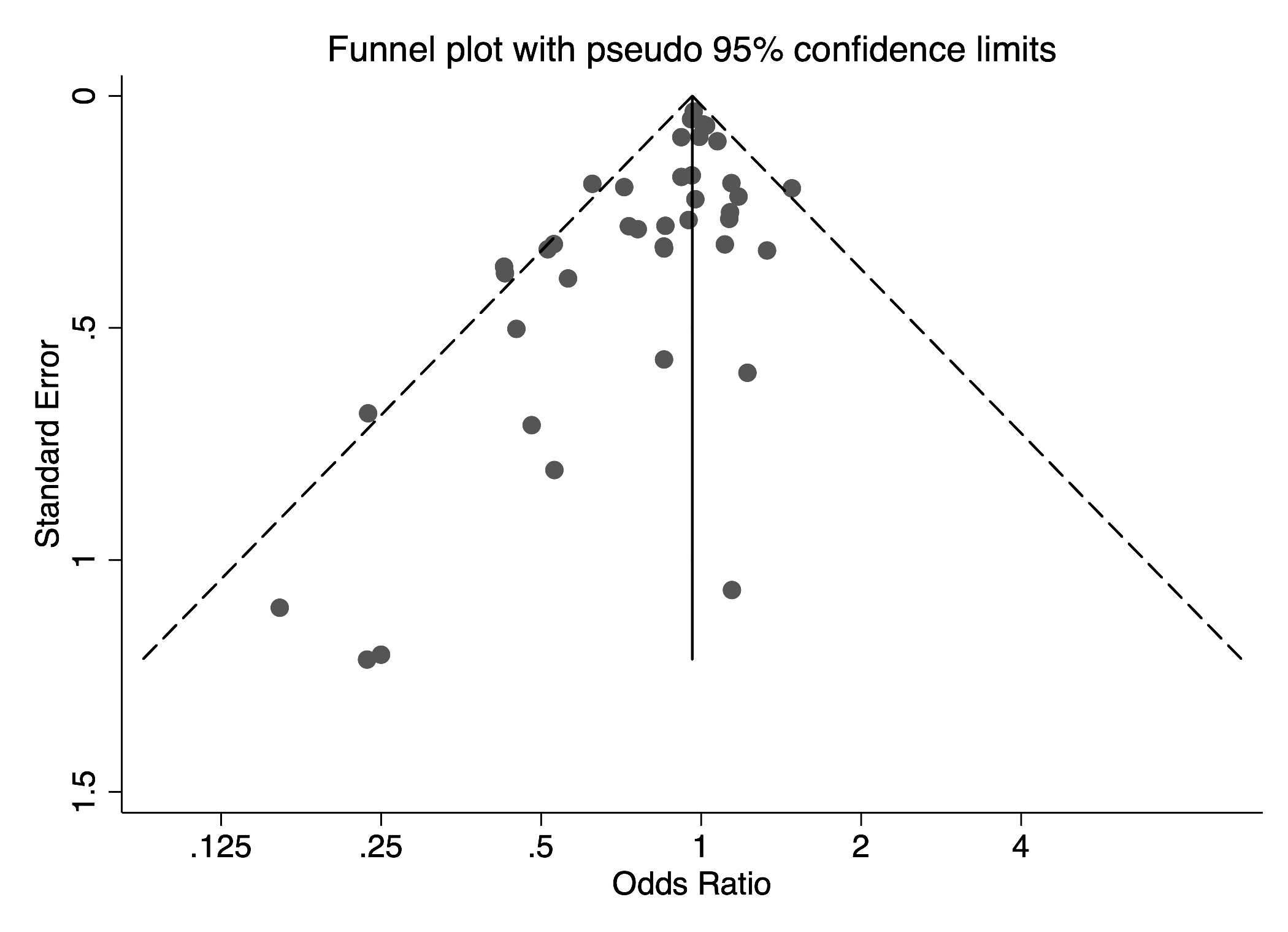

Egger’s test for publication bias: P=0.002

53.	Golan-Tripto I. The Effect of Vitamin D Administration to Premature Infants on Vitamin D Status and Respiratory Morbidity (NCT02404623). <https://clinicaltrials.gov/ct2/show/NCT02404623>.
